# Same answer, different reasons: AI-moderated interviews uncover topics and reasoning that surveys miss

**DOI:** 10.64898/2026.11.01.26367029

**Authors:** Alex Anwyl-Irvine, Marcos Economides, Tomas Engelthaler, Staci Sutermaster, Hannah Kemp, Sema Sgaier, Peter Smittenaar

**Author notes:** Corresponding author: Sema Sgaier.

## Abstract

Surveys can reach large samples but capture only pre-defined answers, with identical responses often concealing very different reasoning. We tested whether AI-moderated interviews capture this reasoning at scale. In a pre-registered, randomized, counterbalanced design, 1,953 US adults from a probability-based panel and an opt-in panel completed three AI-moderated interviews as well as a section with closed-ended and open-text survey items. Topics were extracted from transcripts inductively, without a predefined codebook, and validated by a confirmatory follow-up. Participants endorsed 94% of topics as fair descriptions of what they said. 21–51% of topics from the AI interview had no counterpart in the closed-ended battery, and 17–37 percentage points net coverage over open text answers. Among respondents who gave identical closed-ended responses, their reasoning diverged widely in valence and content. AI-moderated interviews surface reasons that closed-ended and open-text items miss, and show how reasoning differs among people who answer alike.

---

Population research on health attitudes and behaviors uses two main types of instrument. Standardized surveys built on predetermined constructs scale to large samples at low marginal cost, and produce responses that are comparable across people and over time. However, they only capture fixed, pre-defined response options, and identical answers can conceal very different underlying reasoning^1,2^. Open-text items relax that constraint by letting respondents answer in their own words, but responses are typically limited in length and depth. Qualitative interviews, on the other hand, give respondents the ability to speak freely, and interviewers can probe, clarify, and adapt to respondents’ answers. Yet they are time-consuming, costly, and do not scale well, often limiting the generalizability of findings.

Large language models (LLMs) have opened a third option. Their rapid improvement on tasks involving language and reasoning suggests they could conduct fluent, adaptive interviews at scale while analyzing the resulting transcripts automatically^3–5^. One early application has been to improve the measurement properties of standardized surveys, for example by clarifying question wording or prompting respondents to expand on a rating^6^. A more ambitious direction uses LLMs to conduct qualitative-style interviews in their own right^5,7^, and a growing number of commercial platforms now offer such interviews. Early evidence for this approach is encouraging, suggesting that LLM-led interviews can follow good interviewing practice, are received well by respondents, and yield data that captures considerations a fixed survey might miss^5–8^. Yet, rigorous evidence that such LLM-led interviews capture more than standard surveys at scale remains limited.

Existing work leaves four questions of evidence unanswered. First, to our knowledge, no previous study provides a randomized, counterbalanced comparison of AI-moderated interviews against multi-item comprehensive survey batteries, designed to match each subject of the interview prompts. Second, the analysis of AI-moderated interview transcripts has generally relied on researcher-defined codebooks that restrict analysis and cannot, by design, quantify what falls outside them^4,9,10^, reintroducing the very constraints that motivated the interview over a standard survey. Recent work comparing AI systems has begun to surface and quantify topics that emerge beyond a predefined guide, but only in small samples and without comparing to a matched survey^11^. Third, this work has been conducted almost entirely in opt-in panels, with a notable exception^6^, even though it is well established that findings can differ by sampling frame^12,13^. This is a problem because current evaluations of these approaches do not account for sample-noise. Finally, little is known about how the configuration of the interviewer, such as the prompting strategy that governs follow-up questions, affects the number and range of topics surfaced. Early work suggests that different probing strategies are better suited to different contexts, for example idiographic prompts (asking for personal examples) perform better, but this has only been tested with small samples and with probes attached to individual survey questions, rather than in interviews at scale^14^.

This study addresses these gaps in a preregistered, randomized design using a mixed-methods survey platform that allows responses elicited from AI-moderated, multi-turn interviews to be compared directly with standard survey questions. We chose healthcare trust as the test case because trust is contextual, conditional, and shaped by personal experience and political beliefs^1^, This kind of nuanced evaluation is difficult to capture with closed-ended questions. The topic is also timely: trust in US physicians and hospitals fell from around 71% in 2020 to 40% in 2024^15^ yet people still trust individual clinicians more than the system as a whole^16^, and fifty years of trust research has relied heavily on cross-sectional surveys.

Participants provided responses about two healthcare trust domains. Domain 1 examined trust in decisions, covering past experience, and weighing professional advice against social media. Domain 2 examined how trust forms and changes over time, covering how and why trust had shifted in recent years, and what is needed to rebuild it. Each participant completed one domain as an adaptive AI-moderated interview and the other as a standard question battery of closed-ended and open-text items, counterbalanced by domain and order (see Study Design). The battery was built to best-practice standards from published instruments. Interviews were coded using an LLM-based pipeline that extracted labels inductively, repeated ratings, reconciled discrepancies, and clustered the results into a unified topic set^4,17,18^. We fielded the same instrument in two independent samples of US adults, one from a probability-based panel (AmeriSpeak by NORC) and one from an opt-in panel (Prolific), for a combined N = 1,953, allowing us to assess whether AI-derived insights hold when compared to a different sampling method from the same population

This design lets us ask five questions. First, do interviews capture more topics than closed-ended and open-text items? Second, do identical closed-ended ratings conceal different underlying reasoning? Third, as an exploratory question, does the interviewer’s probing strategy (the rules that govern when and how it asks follow-ups) relate to what interviews reveal? Fourth, how do participants experience an AI-moderated interview compared with a standard survey? And fifth, do the topics surfaced hold across probability-based and opt-in sampling frames?

## Results

### Interviews cover topics that matched survey items do not

We needed to derive structure from our interviews, so we applied an inductive topic modeling pipeline to the AI interview transcripts (see Methods) to extract topics, i.e. the reasons underlying participants’ beliefs/behaviors, from transcripts. Each participant’s interview was given a set of labels, each linked to the relevant passage of the interview. We then validated these topics with a sub-sample of 403 participants, who marked 94% of topics as being an acceptable description of what they said or meant Having established that the topic labels extracted from AI-moderated interviews were faithful representations of interview content, we compared our AI interviewer topics to both a typical multiple/single choice battery, and similarly worded open-text box questions. These survey questions were written against the same interview specifications the AI received and were grounded in 17 published instruments and reviewed by two survey methodologists (Methods).

We quantified coverage in two ways. First, against the closed-ended battery, where 21–51% of reliably estimated interview topics (median 19 per subdomain, range 12–28) had no matching item (**Figure 1a**; matching matrix is in the supplement).

**Figure 1.**
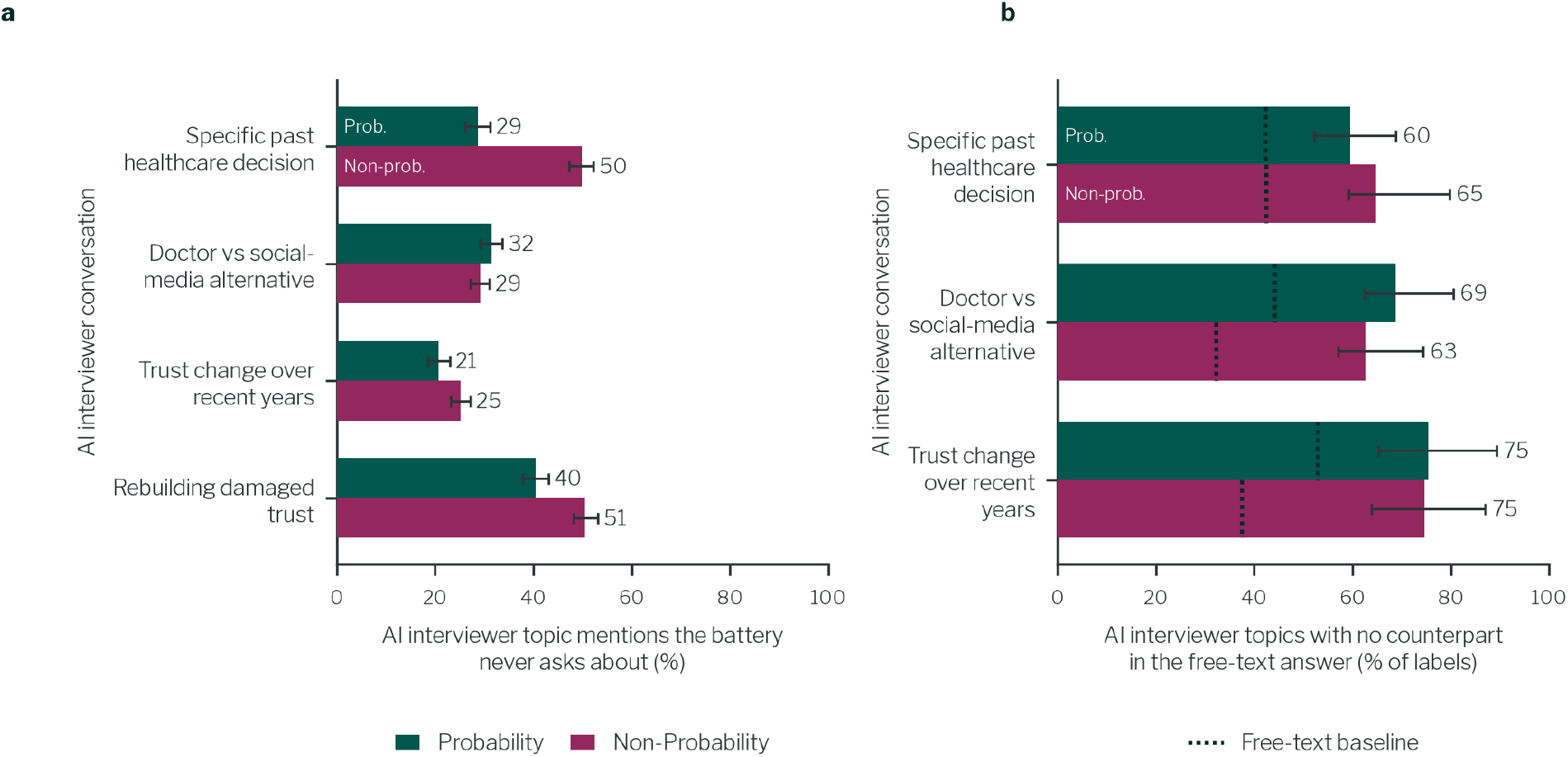
Topics surfaced by the AI interviewer that the standard survey formats do not capture. Each row is one subdomain covered in both an AI interview and a regular survey section. a) Share of AI-interviewer topics that were unique to the AI interview, i.e. lacked a match in the closed-ended battery, by subdomain and sample. Bars are mention-weighted percentages over topics meeting the reliability floor (≥ 12 respondents); error bars are 95% percentile confidence intervals from a bootstrap resampling respondents. b) Share of distinct AI-interviewer topics with no counterpart in the matched free-text answer, by subdomain and sample. Dotted lines show the share of free-text topic labels with no counterpart in the AI interviews (32.3–52.9%). The distance from the dotted line to the end of each bar is the net AI-interviewer difference, accounting for natural heterogeneity across respondents (17.3–37.2 percentage points). Error bars are 95% intervals estimated from both respondent variability and uncertainty in the matching threshold.

Secondly, we applied the identical topic modeling pipeline to open-text boxes which asked the same question as the opening question asked by the AI interviewer. The resulting labels were matched to the interview labels by embedding similarity (see Methods).

Against these matched free-text answers, interview topics had a net advantage of 17–37 percentage points (Figure 1b): 60–75% of interview topics had no counterpart in free text, against 32–53% in the reverse direction (ie. free text topics with no equivalent in the AI interview topics), a reference for how much two groups of people say differently by chance. Both formats surface some unique topics, but the interview consistently surfaces more, and the advantage holds at every matching threshold tested (Methods, Extended Data Fig. 1).

### AI-moderated interviews capture heterogeneity in the underlying reasoning for identical survey responses

We wanted to explore whether AI-moderated interviews could expose different reasoning processes obscured by identical answers in closed-ended batteries. We looked at a single battery which all participants completed after the main study (described in Methods). This focused on trusting different sources of information for health decisions, with each source rated on a likert scale. In this battery, among participants who gave the same closed-ended trust rating, the emotional valence of their interview reasoning ranged widely (interquartile ranges 0.43–1.34 on a −1 to +1 scale; **Figure 2**).

**Figure 2.**
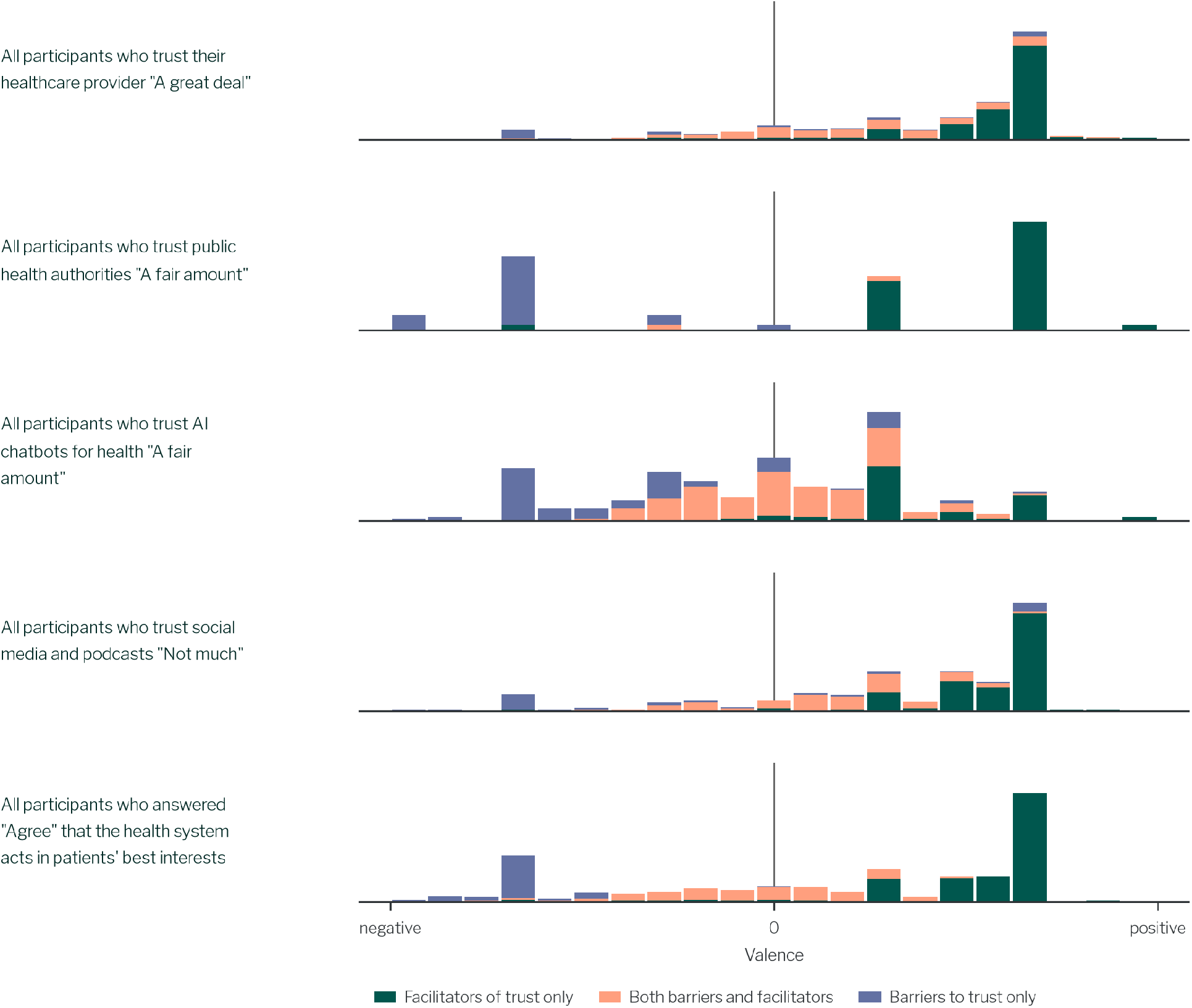
Histograms of respondents showing AI interviews reveal the diversity of reasoning behind a single survey response. Each row shows one battery item and its most common response, pooled across both samples. The x axis is each participant’s mean valence across relevant topics mentioned by that respondent, from −1 (negative) to +1 (positive). Bar height represents the proportion of respondents in that bin. Colors show whether a participant’s relevant topics were facilitators of trust only, barriers to trust only, or both. The interquartile range of valence, top to bottom, is 0.43, 1.34, 0.65, 0.52 and 0.91. Only participants who gave that response and mentioned a topic about the item’s subject in their AI interview are included, from top to bottom: 980 of 1,087; 56 of 842; 306 of 692; 385 of 826; 390 of 865.

As an example, within the group of participants who all said they trusted public health authorities “a fair amount”, the AI interviewer elicited distinct topics and negative opinions about certain bodies. These included concerns about the CDC: “The CDC has become skewed politically, and needs to return to backing what research is actually saying” (topic: Political influence on health agencies, valence=-0.67, topic prevalence 18%), and people distrusting motives due to COVID “We had bad experience with COVID vaccine so it would be difficult to earn my trust” (topic: Distrust of institutional motives, valence=-0.67, topic prevalence 30.4%). The interviews also revealed potential intervention pathways to increase trust: “I look for facts such as clinical studies that give quantitative results” (topic: Scientific evidence validation, valence +0.67, prevalence 37.5%). People who gave the same rating held different, sometimes opposing, reasons, which a closed-ended item cannot show.

### New topics are revealed over multiple turns of the AI interview

We profiled the cumulation of topics over an interview. Across all interviews, extracted topics accumulated steadily as the interview progressed (**Figure 3a**): with every additional turn, participants revealed more unique factors about their beliefs. As the topics identify distinct factors a person covers in their interview, these topics indicate richness, rather than simple words, and their interpretation is endorsed by participants themselves.

**Figure 3.**
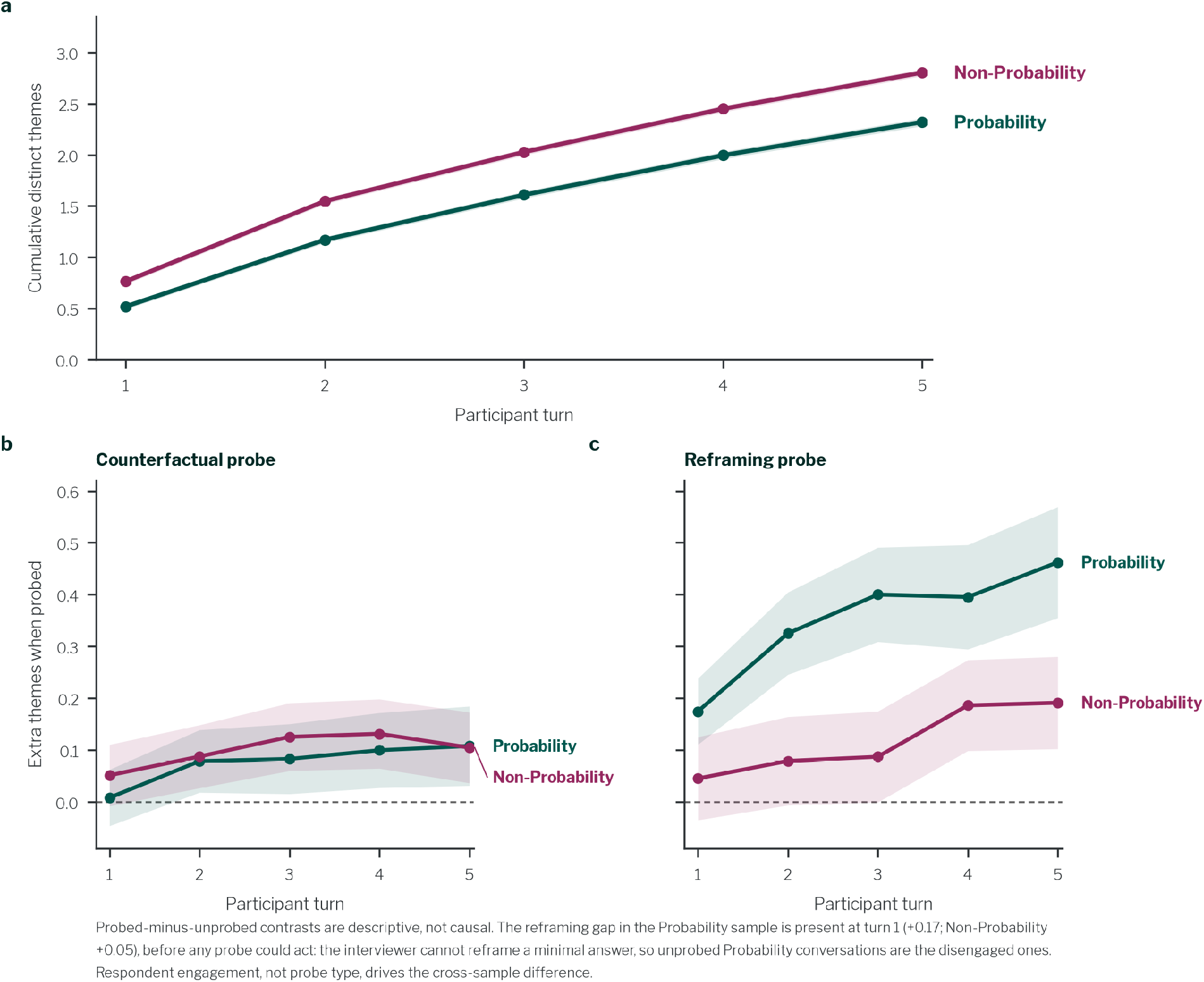
Topics increase across turns of the interview, and two interviewing techniques are associated with additional topics. a) Cumulative topics extracted at each turn, showing a steady but diminishing increase. b) Interviews that used at least one counterfactual probe had slightly more topics; the difference is small but survives correction in both samples. c) Interviews that used at least one reframing probe had more topics in both samples; the association is larger in the probability sample. Shaded areas reflect 95% CI of the difference in average topics between turns using the specified technique and all other conversations.

From first open-response to the final follow up, we see an average of 4.5x more topics in the Probability panel (0.52 =>2.32) and 3.7x (0.77 => 2.81) in the non-probability sample. The returns diminished monotonically, with mean increments of +0.65, +0.44, +0.39 and +0.33 topics across turns 2-5 in the probability sample and +0.78, +0.48, +0.42 and +0.35 in the non-probability sample. Each additional turn therefore yields a smaller, though still positive, return. while the pattern does not asymptote in five turns, it is likely to reach that level at some point.

### Interview probing strategies associated with topics revealed

The AI interviewer had discretion to deploy different probing strategies. We explore two examples: counterfactual (asking what the participant would do under different conditions) and reframing (asking them to reconsider through a different lens). The interviewer chose when to probe, so these are descriptive associations rather than causal estimates.

Counterfactual probes (16–17% of interviewer turns) were associated with about 0.1 more topics from Turn 2 onward in both panels, a small effect (Cliff’s δ ≤ 0.08; Figure 3b, Extended Data Table 1).

Reframing probes (38–42% of turns) were associated with up to half a topic more by Turn 5 in the probability panel and a smaller, later difference in the opt-in panel (Figure 3c, Extended Data Table 1). Interviews that went on to be reframed already held more topics at Turn 1, so the gap partly reflects which conversations the interviewer chose to reframe.

### Differences in user experience between a traditional survey and an AI interview

Being exposed to the AI interview led to some differences in drop out and user experience. In the first section of the survey, where each person had seen only one format, 84.7% of probability-panel respondents completed the AI interview against 95.3% for the battery, a 10.6 percentage point gap (95% CI 7.1–14.0); in the opt-in panel the gap was 7.7 percentage points (Table 1). Multi-turn AI-moderated interviews had not previously been fielded on this panel, which may explain its larger gap.

**Table 1.** User experience outcomes by panel. Drop-out is the completion-rate difference in the first section. The experience composite averages four five-point items; differences are within-response (AI section minus standard section), overall and by section order. Preference was asked after the AI section; the interaction compares the AI-minus-human share between panels. pp, percentage points. ** P < 0.005, *** P < 0.001, Benjamini–Hochberg adjusted.

| Outcome | Probability panel, estimate (95% CI) | Opt-in panel, estimate (95% CI) |
|---|---|---|
| First-block drop-out, AI minus battery (pp) | −10.6 (−14.1, −7.1)<sup>***</sup> | −7.7 (−10.5, −5.0)<sup>***</sup> |
| Experience composite, AI minus battery: all orders | −0.16 (−0.22, −0.10)<sup>***</sup> | −0.06 (−0.11, −0.02) |
| AI block first | −0.06 (−0.13, 0.01) | −0.03 (−0.10, 0.03) |
| AI block second | −0.25 (−0.34, −0.17)<sup>***</sup> | −0.10 (−0.16, −0.04)<sup>**</sup> |
| Future interviewer preference, AI minus human share (pp) | −21.2 (−26.9, −15.4)<sup>***</sup> | +7.1 (0.8, 13.3) |
| Panel × preference interaction (pp) | +28.2 (19.7, 36.7)<sup>***</sup> |  |

Experience ratings were mostly positive (Figure 4a) but slightly lower for the interview than for each participant’s own standard questionnaire section (−0.16 and −0.06 on a five-point scale). The gap was dependent on order: with the interview first, the difference was not significant in either panel; with the interview second, it was (Figure 4b, Table 1). Participants rated the interview less favorably when it came late in the survey.

**Figure 4.**
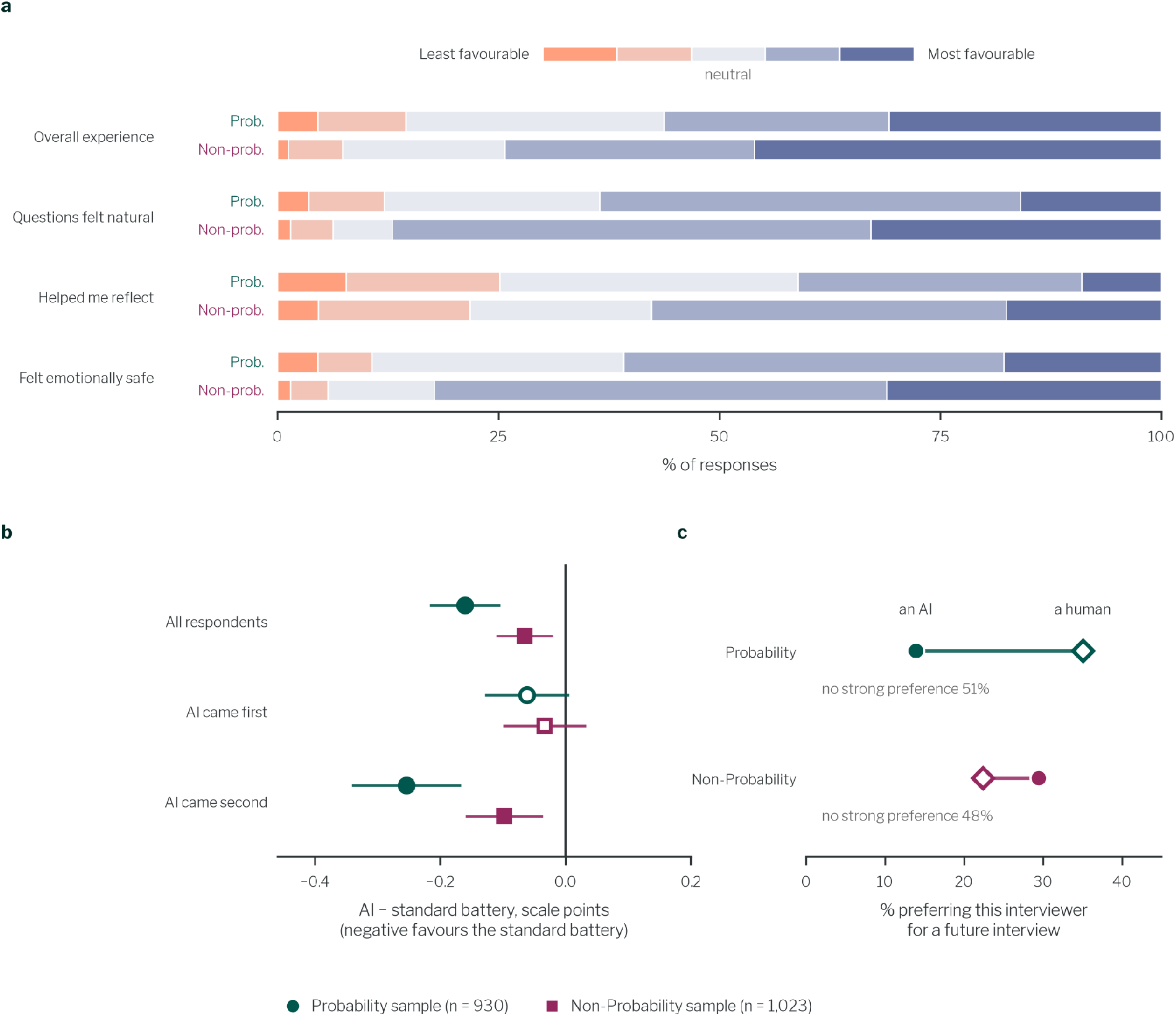
User feedback was mostly favorable, with an order-dependent small preference toward standard questions. a) The distribution of user experience responses after the AI interview, showing the majority across all 4 questions and both panels indicated the experience was positive. The only exception was ‘Helped me reflect’ in the Probability panel. b) Comparing the difference in user experience (averaged across the 4 questions). Questionnaire superiority was only present when the AI was the second section they had done. c) Respondents from the probability panel preferred a human interviewer over AI, whereas the non-probability panel had a slight preference for an AI interviewer.

Asked which interviewer they would prefer in future, about half of respondents in each panel had no preference. Considering the remainder, the probability panel preferred a human (35% vs 14%) and the opt-in panel an AI (30% vs 22%), The AI-minus-human share differed between panels by 28 percentage points (95% CI 20–37; Figure 4c, Table 1): opt-in respondents were more likely to prefer an AI than a human, and probability-panel respondents the reverse.

These AI-related lower ratings are small and order-dependent, but consistent with previous work^6^; we return to it in the Discussion.

### Topic findings replicate across panels

We used the same similarity-based matching approach from our earlier open-text vs AI interviewer comparison to quantify the overlap between the topics identified across the two panels. At the same similarity threshold (*t=*0.7), 59% (58 out of 98) of the Probability panel’s distinct labels and 51% (65 out of 128) of the non-probability panel’s distinct labels had a similar counterpart in the other panel; weighted by how many participants raised each label, coverage reaches **71%** and **61%** of participant-mentions respectively. This overlap is visualized in **Figure 5b**. We also find that the non-probability panel generated more topics. This is likely because they seemed to engage more with the questions (323 vs 162 median words per interview). Accordingly, the non-probability panel covered the probability panel’s topics more completely than the reverse (**Figure 5**).

**Figure 5.**
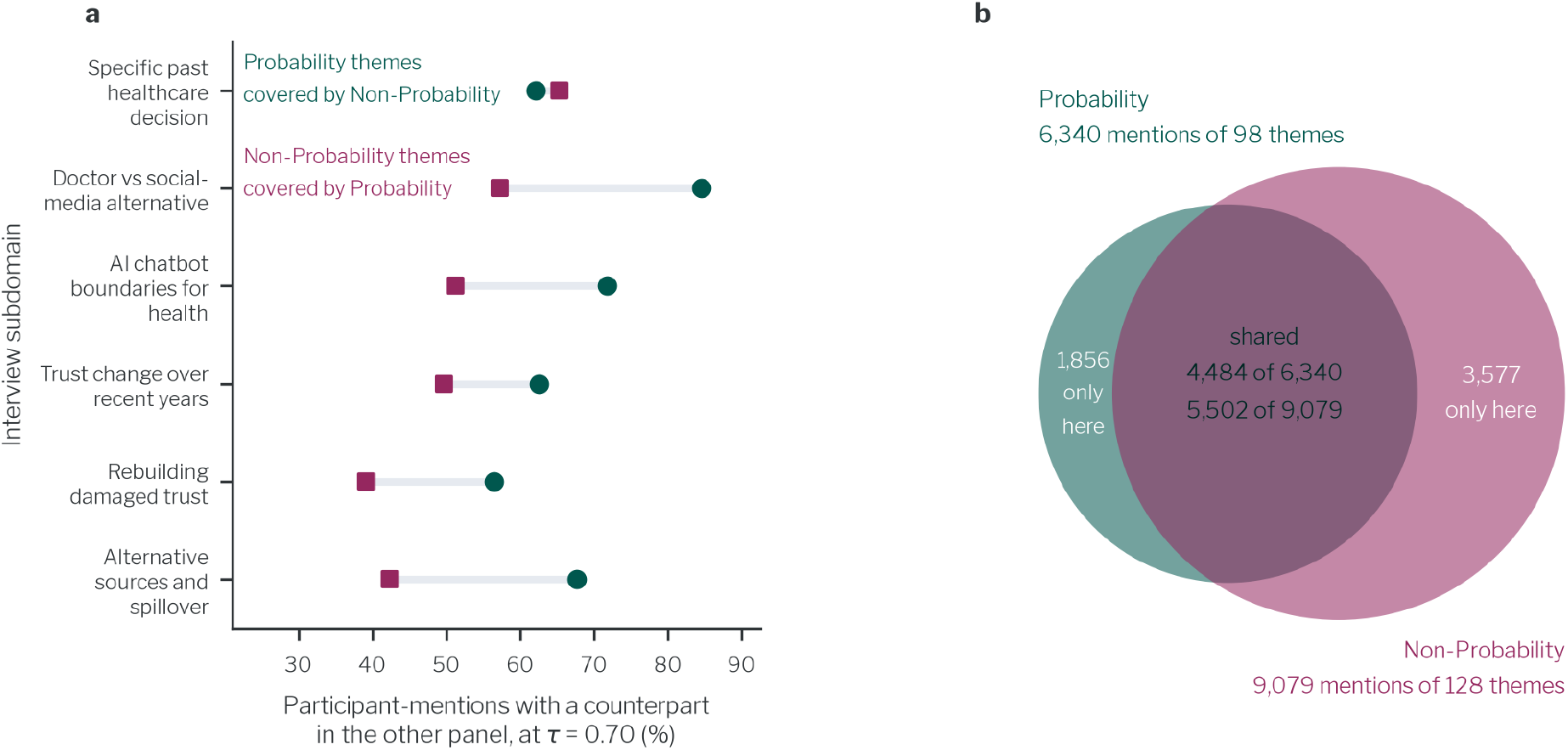
Overlap between the probability and non-probability panels in their topics. a) Coverage for each subdomain, x axis shows the proportion of mentions which have a corresponding label in the other panel, each subdomain has one dot for each panel. Coverage spans from 39-84%. b) Venn diagram shows overlapping and unique mentions of labels across each survey. Pooled, 71% of the probability and 61% of non-probability mention volume is shared.

The sampling noise level associated with survey research across different panels can lead to differences of 8-10pp^19^ in response patterns. We can also expect an online non-probability panel to replicate around 72.5%^20^ of results in an identical study on a representative sample. Taking this into account, our findings here show a reasonable level of overlap (mention-weighted 61-71%)

To further understand how different the panels may be in their perceptions, we also compared the panels on their trust in health information sources (Figure 6). We generally find strong agreement between both panels, with one notable exception: the probability panel is much less trusting of AI (25 percentage point difference). As such, studies exclusively working with non-probability panels may overestimate respondent’s willingness to engage with AI, whether for healthcare or in AI interviews themselves.

**Figure 6.**
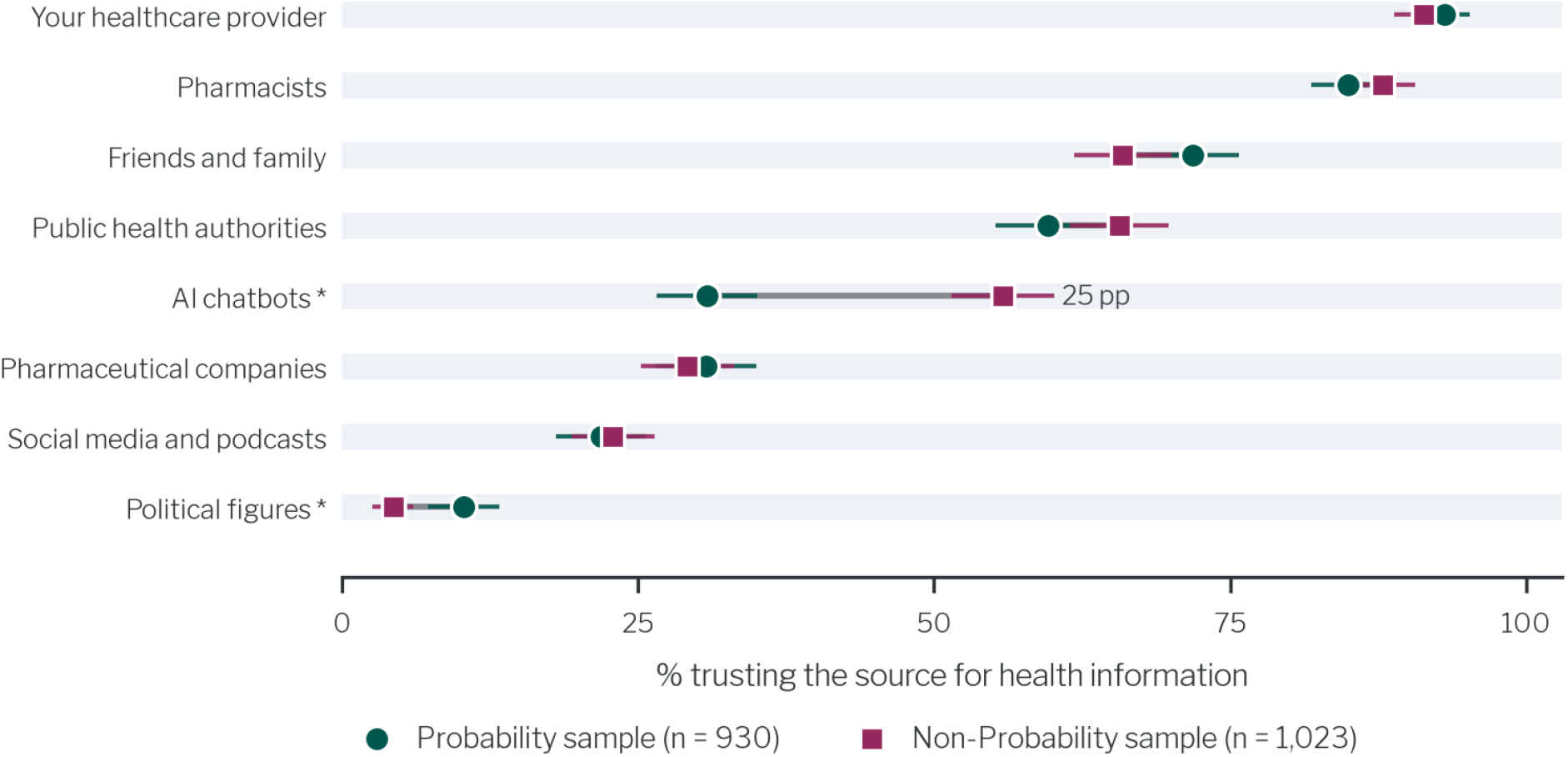
Panels show close alignment on who they trust for health information, with the exception of AI chatbots. The non-probability panel trusts AI chatbots for health 25 percentage points more, and a small ∼6pp difference in political figures. The x-axis shows the proportion of each panel who trust each source ‘A fair amount’ or greater on a likert scale. * indicate points which were significantly different between the panels.

## Discussion

We show that AI-moderated interviews can surface reasoning that matched survey items miss. We were able to capture individual reasoning patterns that surveys often fail to capture while retaining the scale and reach. Specifically, we show that across subdomains, between 21% and 51% of interview topics from the interviews had no equivalent in the matched closed-ended survey, and that we could identify these inductively, without a predefined codebook.

The coverage findings were consistent in both a probability-based panel and an opt-in panel of US adults, to our knowledge the first test of an AI-moderated interviewing method in independent probability-based and opt-in samples. This matters because sample source is known to affect questionnaire and survey outcomes, and because most work in this area has been conducted on a single opt-in panel^5–8^. The main divergence between panels was greater distrust of AI and greater dropout during the AI interview section in the probability panel, which is worth noting for researchers working in this area, since it suggests panel choice may shape findings on AI-related questions more than on other domains.

Our findings on trust largely replicate previous findings from the literature, such as participants trusting their own clinician more than the health system as a whole, and reporting a general erosion of trust since the pandemic. But the AI-moderated interviews were able to surface some of the specific individual experiences driving those patterns. For example, participants who gave the same closed-ended trust rating (trusting public health authorities “a fair amount”) described very different experiences behind this, and their reasoning spanned barriers and facilitators of trust. A survey would place these people in the same group, yet they hold different underlying attitudes and it is possible these people would respond to attempted interventions in a different way.

Interestingly, we also found that the probing strategy adopted by the AI interviewer was associated with how much information surfaced. Specifically, turns using a counterfactual or reframing probing strategy were associated with more topics being surfaced, though this was observational rather than experimental in the sense that the AI interviewer chose when to deploy each technique. This suggests that interviewer design plausibly affects what an interview can uncover, consistent with theoretical accounts of why probing deepens qualitative data^21^, though establishing a causal effect requires further controlled studies. It also suggests AI interviewing tools should not be treated as interchangeable: how an AI interviewer is configured may have substantial impact on the quality of the results.

These findings could have wide application across several settings. Qualitative research is expensive, slow, and limited by the availability of trained interviewers. An AI interviewer running over the web or by phone could collect open, probed accounts at survey scale on questions like vaccine hesitancy, care-seeking, or maternal health, and could work across languages without recruiting and training local interviewer teams. For health systems trying to improve patient experience, this method could recover the nuance behind closed-ended ratings for an entire patient population, rather than just from small numbers recruited into focus groups. Similarly, in pharmaceutical and market research, this method could help to uncover the reasoning behind important behaviors such as treatment adherence and trial participation, which are usually only studied through small qualitative samples, and which might not generalize to larger populations. In public health and policy, attitudes can change faster than validated instruments can be built. For example, attitudes towards vaccination shifted rapidly during the COVID-19 pandemic, and a similar pattern can be seen today with regards to the use of AI in healthcare. Since our topic extraction method does not rely on a predefined codebook, it allows novel topics to be studied and reported without having to fully anticipate those topics in advance.

There are several important limitations of our study. First, what we compare against. This study did not compare AI interviews against human-led interviews, and further work is needed to test how closely these insights match human-level depth. Our comparison is based on an instrument that was designed for this study. We designed the quantitative sections using a careful process of literature review, evaluation of validated scales, and thorough testing. The domains did not allow exclusive use of psychometrically validated scales, so coverage figures come from our instrument rather than fixed and purely validated properties. A longer battery might have captured more of what the AI interviews surfaced.The closed-ended comparison also covers four of six subdomains and the open-text comparison three (due to design constraints detailed in the Methods). Finally, modality was compared between subjects to avoid contamination, so the comparison rests on randomization rather than paired within-person data. This choice is supported by recent experimental evidence that answering AI interview probes changes subsequent closed-ended responses^22^.

Secondly, there are limitations related to how topics were generated, validated, and used. The interview model, our prompting scaffold, and our analytics pipeline may have limited the topics that surfaced. Because our member checking sub-study showed participants only their own topics, we cannot separate accurate labeling from acquiescence. Previous labeling confirmation has shown this only leads to a slight false positive rate^6^, but we flag this as a limitation. Foil topics from other participants would separate the two. Finally, the association between probing style and topic coverage is observational because the interviewer chose when to probe, so we cannot separate greater engagement leading to more labels against the method leading to more labels.

Thirdly, there are limitations regarding participants and generalizability. Even though people rated the AI interviews as acceptable, we did see a cost, higher early drop-out, slightly lower experience ratings, and a preference for human interviewers in the probability panel. This pattern matches prior work^6^, which also found higher drop-out and slightly lower ratings for AI-moderated follow-ups. The impact is small and order-dependent: ratings fell only when the interview came second, so part of it reflects fatigue late in the survey rather than the format itself. We interpret this as a trade-off, a small loss in experience against a substantial gain in topic coverage, and one that improvements to the interviewing interface are likely to narrow. Lastly, the study covers one subject area, trust in healthcare, in one country. while this is a broad domain, touching health, trust and politics, other subjects may vary in findings on insights and overlap.

Future work could address some of these limitations and further strengthen support for AI interviewers. For instance, Experimentally manipulating probing strategies, validating topics against foil labels and human coders, and testing other subject areas and countries, would improve evidence. And because interviewer and analysis models are improving quickly, we expect the capabilities to increase and evaluations like this one should be repeated as the tools change rather than treated as fixed.

Our findings show that AI-moderated interviews can reveal a substantial amount of the reasoning that standard surveys leave uncaptured, at survey scale. Across public health, health services, and clinical research, the reasoning behind a response has rarely been measured in large samples, despite being crucial to identifying an intervention target, or a change to a protocol or policy. Recovering such reasoning in large samples could substantially extend what survey research is able to deliver in these settings

## Methods

### AI-moderated Interviewer engine

The AI interview was executed based on a configuration set up by the authors. The configuration fully specified the sequence of quantitative and AI-driven questions for a given arm, and four such configurations were created to serve the four arms (Extended Data Fig. 2). It also specified an overall set of instructions for the role the AI was to play, akin to a “system prompt” in other AI products. Finally, it specified which content safeguards were active to stop the interview for participant safety, specifically we used Azure AI Content Safety, which flagged conversations covering topics like self-harm & violence. As people were asked to speak about sensitive topics we applied a higher threshold of 6 on Azure’s eight-level severity scale (0–7). We set this after manual testing, to reduce false-positive interview terminations found at lower thresholds. The model used in this configuration was OpenAI’s gpt-5.4-mini-001.

For the AI-moderated interview sections - of which each participant completed three (Extended Data Fig. 2) - we specified the objective of the interview, the number of turns available to the AI, the large language model endpoint to use, the fixed opening question, what context from previous questions should be considered, and few-shot examples of good and bad follow-up questions identified during testing. When a respondent answered the opening question, the LLM was prompted with all the aforementioned information, and a followup question was formulated and transmitted to the web interface.

### Study design

#### Sample and recruitment

The instrument was fielded to two independent samples of US adults aged 18 and over, drawn from panels with deliberately different recruitment logics so that the findings could be tested against a sampling frame.

The probability sample was collected using a nationally representative sample drawn from the AmeriSpeak Panel, a probability-based panel operated by NORC at the University of Chicago. This panel is recruited using a rigorous multi-mode strategy (mail, telephone, and face-to-face non-response follow-up), to ensure comprehensive U.S. household coverage. Panel recruitment is by invitation to a sampled address only, with no self-nomination into the panel, so selection probabilities are known and the resulting sample supports design-based inference to the US adult population. Our sample was drawn from this panel using sampling strata based on age, race and ethnicity, education, and gender.

The non-probability sample was recruited via Prolific, an online participant pool in which adults enroll themselves and opt into individual studies. To ensure our findings were generalizable to the broader United States population, we utilized Prolific’s “Representative Sample” allocation feature. This methodology employs a cross-stratified quota sampling approach designed to mirror national demographic distributions based on U.S. Census Bureau data. Specifically, the sampling frame was stratified across a narrower set of factors: sex, age bracket, and ethnicity (using simplified census categories). Prolific is stratified from a self-selected pool rather than a probability sample. Throughout the paper the two samples are labeled Probability and Non-probability, are fielded with the identical instrument, and followed the same randomization rules.

The analyzed sample sizes were 930, and 1023 for the probability and non-probability panels respectively. The studies overrecruited to account for dropouts within the survey, 304 (probability) and 246 (non probability) were not included in the final data set for this reason. We also checked for more than 5 copy-past events (as a proxy for copying from an AI-chatbot), 4 (probability) and 39 (non-probability) were not included for this reason. Demographic attributes of both samples are given in Extended Data Table 2.

**Extended Data Table 2.** Demographic attributes across the two samples. Values are unweighted n (column %) for the analyzed probability (AmeriSpeak, n = 930) and non-probability (Prolific, n = 1,023) samples.

| Attribute | Probability | Non-probability |
|---|---|---|
| Age - Mean (SD) | 48.7 (18.1) | 45.7 (15.9) |
| **Sex** |  |  |
| Female | 421 (45.3%) | 519 (50.7%) |
| Male | 506 (54.4%) | 499 (48.8%) |
| Not reported | 3 (0.3%) | 5 (0.5%) |
| **Race and ethnicity** |  |  |
| Asian or Pacific Islander, non-Hispanic | 43 (4.6%) | 66 (6.5%) |
| Black, non-Hispanic | 125 (13.4%) | 132 (12.9%) |
| Hispanic | 139 (14.9%) | 106 (10.4%) |
| Other, non-Hispanic | 9 (1.0%) | 18 (1.8%) |
| Two or more races, non-Hispanic | 34 (3.7%) | 39 (3.8%) |
| White, non-Hispanic | 577 (62.0%) | 656 (64.1%) |
| Not reported | 3 (0.3%) | 0 (0.0%) |
| Prefer not to answer | 0 (0.0%) | 6 (0.6%) |
| **Education** |  |  |
| Less than high school | 69 (7.4%) | 9 (0.9%) |
| High school graduate | 166 (17.8%) | 118 (11.5%) |
| Some college or associate degree | 336 (36.1%) | 284 (27.8%) |
| Bachelor's degree | 201 (21.6%) | 418 (40.9%) |
| Postgraduate degree | 155 (16.7%) | 190 (18.6%) |
| Not reported | 3 (0.3%) | 0 (0.0%) |
| Prefer not to answer | 0 (0.0%) | 4 (0.4%) |
| **Household income** |  |  |
| <\$20,000 | 104 (11.2%) | 63 (6.2%) |
| \$20,000-\$39,999 | 130 (14.0%) | 155 (15.2%) |
| \$40,000-\$74,999 | 241 (25.9%) | 302 (29.5%) |
| \$75,000-\$99,999 | 128 (13.8%) | 154 (15.1%) |
| \$100,000-\$149,999 | 159 (17.1%) | 212 (20.7%) |
| \$150,000+ | 165 (17.7%) | 115 (11.2%) |
| Not reported | 3 (0.3%) | 22 (2.2%) |
| **Employment status** |  |  |
| Employed (including self-employed) | 571 (61.4%) | 723 (70.7%) |
| Not reported | 3 (0.3%) | 14 (1.4%) |
| Other not working | 41 (4.4%) | 81 (7.9%) |
| Retired | 208 (22.4%) | 118 (11.5%) |
| Unable to work | 63 (6.8%) | 27 (2.6%) |
| Unemployed or laid off | 44 (4.7%) | 60 (5.9%) |
| **Marital status** |  |  |
| Married | 462 (49.7%) | 418 (40.9%) |
| Never married | 299 (32.2%) | 322 (31.5%) |
| Formerly married (divorced/separated/widowed) | 166 (17.8%) | 160 (15.6%) |
| Living with partner, not married | – | 113 (11.0%) |
| Not reported / Prefer not to answer | 3 (0.3%) | 10 (1.0%) |

#### Weighting

The AmeriSpeak panel provided participant weights, which are raked^23^ to US adult benchmarks on age, sex, education, race and ethnicity, housing tenure, household telephone status, Census division, and the interactions age × sex, age × race and ethnicity.

Prolific supplied no weights, so we raked the sample by iterative proportional fitting^24^ to the weighted margins of the probability panel on the same dimensions, omitting household telephone status, which Prolific does not collect, and trimmed the resulting weights to the range 0.2–6.0. Calibrating to the weighted probability sample puts both samples on the same target population without introducing a second external benchmark. Within each panel, weights were rescaled to a mean of 1 over analyzed respondents.

Weights were applied only where we estimated population quantity: the experience ratings, the within-respondent comparisons of AI-moderated and standard blocks, the interviewer-preference shares (Figure 4), and the trust-in-sources battery (Figure 6).

Weighted proportions and means are ratio estimates, ∑*_i_ w_i_y_i_*/ ∑*_i_ w_i_*, with linearized standard errors, Wald 95% confidence intervals, and Kish’s effective sample size^23^.

#### Main study

**Extended Data Fig. 2.**
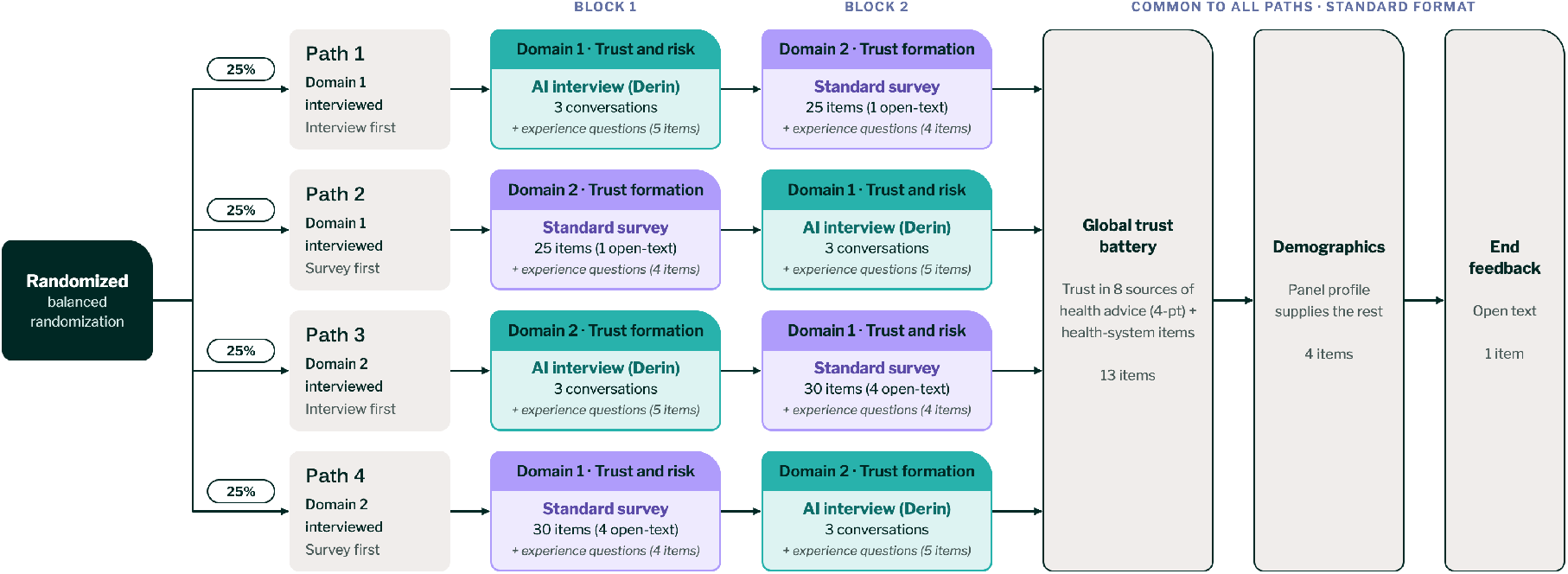
Randomized instrument paths. Participants were randomly assigned to one of four paths. The paths varied which domain was covered as an AI-moderated interview and whether the interview or survey block came first. Each participant completed both domains, one in each format, so format varied within participants and no participant answered the same domain in both formats. Each block ended with experience questions (four items, and an AI-vs-human preference item after an interview block). All paths then finished with the same standard-format trust battery, demographic items and open-text feedback.

Participants were allocated to one of four paths illustrated in **Extended Data Fig. 2**. They completed two domains on trust in healthcare: Domain 1, trust and risk trade-offs in specific decisions; Domain 2, personal history and trust formation. One as a block of three AI-moderated interviews and the other as a standard block of closed-ended and open-text survey items. Each block was followed by an experience questionnaire.

Which domain was AI-interviewed and which block came first were randomized jointly, giving four paths (interview-first or survey-first × Domain 1 or Domain 2 interviewed). No participant had the same domain in both formats, so our comparisons of content coverage between the modalities account for subject.

Each interviewed domain comprised three interviews, giving six interview subdomains across the two domains. Domain 1 covered a specific past healthcare decision in which the respondent followed or rejected guidance (labeled T1-1), weighing a doctor’s advice against a less-proven alternative promoted on social media (T1-2), and where the respondent draws the line on using AI chatbots for health questions (T1-3). Domain 2 covered how and why the respondent’s trust has changed in recent years (T2-1), what the healthcare system would need to do to rebuild trust (T2-2), and alternative information sources and the spillover of trust between them (T2-3). Each interview opened with a scripted question and then ran for up to five participant turns, with the interviewer choosing its own follow-up at each turn from the specification for that subdomain. Two subdomains (T1-1 and T2-1) are gated: the opening question is a yes/no branch and a respondent sees exactly one branch, which we treat as a single subdomain in analysis.

Each block was followed by the same four-item experience questionnaire: overall experience, whether the questions felt natural and relevant, whether the section helped the respondent reflect on things they had not thought about before, and whether they felt emotionally safe sharing sensitive information. This was designed so that experience is measured within a person across the two modalities. A fifth item, asking whether the respondent would prefer this kind of interview to be led by an AI or by a human in future, was asked only after an AI interview block.

Participants were lastly shown the same closed-ended question block before randomization: a trust-source battery asking how much they trust each of eight sources of health recommendations (their healthcare provider; pharmacists; people close to them; public health authorities; pharmaceutical companies; AI chatbots; people they follow on social media or podcasts; political figures) on a four-point scale, together with a single agreement item on the healthcare system.

Of the six interview subdomains, four had a matched closed-ended battery (T1-1, T1-2, T2-1, T2-2). Two subdomains, trust in AI for health (T1-3) and alternative information sources (T2-3), were added to the AI interview arms only, after feedback during instrument design identified them as timely; we did not add matching battery items, to limit respondent burden. Three subdomains (T1-1, T1-2, T2-1) also had a matched open-text item. Rebuilding damaged trust (T2-2) did not, so it appears in Figure 1a but not Figure 1b.

### Instrument Design

Instrument development involved three stages, which were domain selection, interview specification, and item writing. Domains were selected on the basis of a narrative review of research and media coverage on trust in healthcare in the United States published between 2019 and 2026, drawing on approximately 30 academic and policy sources alongside national news reporting. The review asked where trust in healthcare currently stands, how it varies across people and contexts, what drives it up or down, and how academic and media accounts of these trends diverge. It identified four areas the literature has not resolved. These are how trust is repaired after it erodes, why people switch health information sources and how they justify switching, how risk calculus interacts with trust when people follow health guidance they doubt or reject guidance they trust, and how trust operates where human and AI sources are combined. Two domains were selected out of the four: trust and risk trade-offs, and personal history and trust formation. Both were selected because there are many potential reasons how people arrive at their current beliefs and understanding the reasoning in more depth is valuable. The initial literature and media scan was generated using ChatGPT Deep Research in January 2026. All findings were subsequently reviewed and cross-checked against a selection of systematic reviews and media sources by the authors, who produced the final synthesis and domain recommendations.

The interviews were specified next, setting out what each interview should cover and probe, and the survey items were derived from those specifications. Each interview aim was translated into one or more closed-ended or open-text items targeting the same content. Where an aim covered several distinct ideas, a battery was written rather than a single item, so the survey had room to cover necessary ground albeit within the constraints of closed question types.

To ground the items in existing measurement, 17 published instruments, national survey programs, and theoretical frameworks were reviewed. These were the Health Care System Distrust Scale (HCSD) and its revision, the Medical Mistrust Index (MMI), the Medical Mistrust Scale for Medical and Healthcare Students and Providers (MMtS-MHC-SP), the Trust in Physician Scale (TPS), the Wake Forest Physician Trust Scale short form (WFPTS), Trust in the Medical Profession (TMP), the Trust in Multidimensional Healthcare Systems Scale (TIMHSS), Trust in Public Health Authorities (TiPHA), the Decisional Conflict Scale (DCS), shared decision-making scales (e.g., SDM-Q), the Autonomy Preference Index (API), the Multidimensional Health Locus of Control Scale (MHLC), the Theory of Planned Behavior, the Ipsos PX Pulse, the Health Information National Trends Survey (HINTS), the KFF Health Tracking Poll, and the NORC/ABIM Foundation trust survey. Where possible, items were adapted from these scales, or written against constructs those scales define. To maximize equivalence between the two arms, however, most items were constructed new, since established measures did not cover the domains or framing that a fair comparison required.

An example of an adapted item is “Important medical decisions should be made by the doctor, not the patient”, adapted from the API (original: “The important medical decisions should be made by your doctor, not you”), and an example of an item based on a validated construct is “I distrust the healthcare system overall, but still trust a doctor”, written to capture the distinction between system-level and provider-level trust that the HCSD and the TPS treat separately. Batteries were largely constructed from mixed sources. For example, in the battery asking what drove uncertainty in a past health decision, the four statements correspond respectively to competence, drawing on TiPHA and TIMHSS; benevolence, drawing on the HCSD; risk and uncertainty, drawing on the DCS; and normative pressure, drawing on shared decision-making scales and the Theory of Planned Behavior.

Response formats followed standard conventions. Closed-ended items used four-point Likert or single-choice formats. Rating batteries were paired with a forced-choice follow-up asking which factor mattered most, thus distinguishing endorsement from priority. Critically, we added open-text items where relevant, as these are a standard part of traditional surveys, and allowed analysis of what a multi-turn interview adds over a single open-text response.

The instrument was reviewed internally by two coauthors with extensive experience in quantitative survey design, who assessed wording, response format, and coverage against the interview specifications. Three quality assurance pilots were run in March 2026, with 10, 25, and 50 participants, prior to full fielding.These were used to review the quality of the interviews and to inspect survey responses descriptively. No formal analysis was conducted on the pilot data.

### Ethics

The AmeriSpeak sample study arm was reviewed and approved by the Institutional Review Board of NORC at the University of Chicago.

The Prolific arm was reviewed and approved by Salus IRB as Sub-study 2 of Surgo Health’s umbrella protocol 0200, “Understanding Drivers of Health Decisions Across Populations” (Salus reference 26406).

All participants gave informed consent electronically. Panel members consent to research participation on joining AmeriSpeak or Prolific; before starting this study, all participants were told it included an interview with a conversational AI and agreed to a study-specific consent screen on the Derin platform. AmeriSpeak panelists received NORC’s standard survey incentive (equivalent to $5); Prolific participants were paid at a rate of $7 per hour.

### Preregistration

The probability-sample study was preregistered on the Open Science Framework before data collection (OSF Registries 6tzsq, https://osf.io/6tzsq, registered 16th April 2026). The opt-in sample was not registered and was fielded with the identical instrument as a replication. Of four registered hypotheses, H2 (the interview surfaces reasons the closed-ended battery does not ask about) is tested in Figure 1 and supported, and H4 (superior participant experience in the interview format) is tested in Figure 4 and not supported. H1 (more elaborated responses) and H3 (clearer prioritization of reasons) were not evaluated here. Figures 2, 3b,c, 5 and 6 and the member-checking exercise were not preregistered and are exploratory; Figure 3a was pre-specified as exploratory.

We report some deviations from our preregistration:

- Our preregistered plan used participant-level mixed-effects models, with topic and order as covariates. However, the experience outcomes were instead analyzed with weighted mean differences and Wald z tests, and this type of model was not applicable to the coverage analysis reported.
- The coverage analyses treat the topic label as the unit and report bootstrap intervals, because the registered quantity is a property of the set of reasons surfaced rather than a per-participant score, and drop-out is compared with two-proportion tests.
- The registered completion-time exclusion (median − 2 × MAD or five minutes) was applied, though nobody fell into this bin.
- The probability sample closed at 930 analyzable respondents against a target of 1,000; the decision to stop was made prior to reviewing any of the results.

### Analytics

#### Topic labeling extraction

Interview transcripts were converted into per-participant topic labels using a four-stage LLM pipeline, combining concepts from the LLM-evaluation literature on grading rubrics^25,26^, and modern conversation topic harmonization tools like Athropic’s CLIO^17^

1. independent multi-rater extraction
2. consensus adjudication
3. embedding-based harmonization of labels into a per-question vocabulary of topic families
4. generation of a short description for each family.

These steps are detailed below.

##### Stage 1: Multi-rater label extraction

Each transcript was coded independently by three LLM raters (Claude Sonnet 4.6) using the same structured content-analysis rubric (Supplementary Note 1). Single rubric ‘autograders’ have been shown to be the most reliable approach for this type of extraction^26^. The rubric defined the target construct as a topic, i.e. an underlying reason that explains why the participant holds a belief or makes a decision, and required topics to be grounded inductively in the participant’s own words rather than in a predefined codebook. For each topic an LLM rater returned: a lowercase label of at most five words; a one-sentence belief statement; an effect on the target behavior (barrier, facilitator, mixed, unclear); an importance score from 0 to 1, representing the importance of that topic to the person based on the transcript; a valence score on a seven-point scale from −1 to +1 capturing the participant’s feelings toward the topic; the User turn indices where it appeared; and one or two verbatim supporting quotes. Rank was derived from importance rather than assigned independently.

##### Stage 2: Consensus adjudication

A fourth LLM call (Claude Sonnet 4.6) acted as consensus grader^25^ over the three rater outputs for each transcript (Supplementary Note 2). A topic was retained only if at least two of the three raters identified the same underlying reason, allowing close synonyms. Synonymous labels were merged to one canonical label, importance and valence were averaged across agreeing raters. The averaged valence was then rounded to the nearest point of the seven-point rubric scale (−1 to +1), and averaged importance was retained as a continuous 0–1 score. Quotes and turn indices were unioned and deduplicated, and rank was recomputed from importance. The adjudicated output for each transcript formed the raw topic set carried into harmonization.

##### Stage 3: Label harmonization across participants

Raw labels vary in wording across participants, so labels were harmonized into a compact vocabulary of topic families. This was adapted from the method used in Anthropic’s CLIO tool^17^. Harmonization was run strictly per question, so a label string occurring under two questions was harmonized independently in each.

Unique raw labels were embedded with Cohere embed-v3-multilingual. Embeddings were L2-normalized, reduced with UMAP, and clustered with HDBSCAN (settings in Table M1). HDBSCAN noise points were reassigned to their best cluster only when their soft membership probability met a minimum threshold; otherwise they were carried forward as their own candidate so that rare but distinct topics were not absorbed into unrelated clusters.

Clusters were then canonicalized by an LLM judge (Claude Opus 4.7) in three passes (Supplementary Note 3). In Pass 1, labels within each cluster were presented in chunks alongside their frequency and up to two example belief statements and one quote, and the judge mapped every label to a canonical label, merging only labels expressing the same underlying reason. In Pass 2, the per-chunk candidates within a cluster were consolidated to a small set. In Pass 3, candidates from all clusters and unassigned noise labels were pooled with cumulative counts and collapsed into a final family vocabulary with a target of about 20 families. Before Pass 3, near-duplicate candidates were pre-merged by embedding cosine similarity, and large pools were consolidated in chunks followed by a final merge. Every judge prompt forbade collapsing labels into vague umbrellas (for example “concerns” or the subdomain itself) and required family labels to remain specific reasons and at most five words. In practice, some conversations still had un-merged small number labels, we chose to leave these as distinct but uncommon topics that are qualitatively interesting.

The output was a per-question mapping from each raw label to a harmonized family, with provenance (cluster id, noise status, assignment probability) and two diagnostic scores: each raw label’s cosine similarity to its family centroid, and to the embedded family label.

##### Stage 4: Family descriptions

For each harmonized family, an LLM (Claude Sonnet 4.6, temperature 0.3) wrote a one-to two-sentence neutral description synthesized from its child labels, belief statements, and quotes (Supplementary Note 4). These descriptions were used for reporting and were shown to participants in the member-checking exercise.

##### Final output

Each participant-question pair - i.e. a single turn in an AI interview - was now enriched with a list of topics, each with its raw label, harmonized family label, belief statement, effect on target, importance, valence, turn indices, and validated quotes.

#### Validation of topic labels

The labels are crucial to claims about the subject coverage and panel comparison. We conducted a follow up exercise with 403 participants from the non-probability panel. We targeted 100 participants from each study arm (actual numbers per arm 101-105), to ensure the interview subject did not confound the follow-up results.

Participants judged their own answers against one topic at a time and answered two questions about each: whether the label was “a fair description of something you said or meant” (binary) and “how accurately does it capture what you meant” (1 = not at all accurate to 10 = completely accurate). The accuracy rating was requested whether or not the participant endorsed the topic.

Each participant had completed 3 interviews so across the 403 samples there were 4,102 topic label ratings, with a mean of 10.2 labels each (range 1–18). Ratings were clustered within participants, so we calculated confidence intervals with a participant-level cluster bootstrap with 5,000 iterations, whereby ratings were resampled within each respondent.

Participants marked 94% of topics as being an acceptable description of what they said or meant, and rated the accuracy of topic descriptions an average of 8.7/10 (95% CI 8.6-8.8). 77.8% (95% CI 73.5–81.6) of the 401 debrief respondents selected only “nothing missing” when asked whether anything they had wanted to say was absent from the topics.

This value may be inflated by acquiescence, the tendency to agree with statements put to respondents^27^. However, we do see some level of rejection (5.9% of topics), and accuracy ratings variability (median 9 of 10, 49% of topics at the maximum), and previous work shows acquiescence is relatively small in participant evaluation of AI labels^6^.

#### AI interview vs. standard overlap quantification

##### Closed-ended questions

To capture the overlap between closed-ended standard questions and the labels extracted from AI interviews (Figure 1) we built a mapping table of labels to each possible closed-ended answer.

For each label we classified it by LLM as:

Covered: an in-scope label judged to map onto a battery item from the matched subdomain for that question;
Unanticipated: an in-scope label judged not to map onto any battery item in that subdomain (the plotted quantity in Figure 1);
Not evaluated: a label below the reliability floor and excluded

The LLM was given the list of items and response options in the matching battery, and the topic label, and told to classify each alongside a rationale.

This matching table was then reviewed and corrected by an author for accuracy.

##### Calculating the reliability floor

We included a label as a candidate for coverage only when its prevalence within a subdomain can be considered reliable for that sample size. We excluded any labels present for <12 respondents, based on a requirement to have a relative standard error ≤ 30%, the conventional threshold below which a survey proportion is treated as reliable in US federal health statistics^28^

##### Calculating mention-weighted overlap

The number of times a label comes up across participants is important, because labels differ widely in how much of the conversation they represent. Counting labels equally would give one raised by 12 people the same weight as one raised by 200. The mention-weighted share reflects what fraction of participants’ total set of labels concerns something the closed-ended battery does not ask. A mention is defined as a respondent raising one label within a subdomain; a label raised by the same respondent on several turns counts once.

Across the eight combinations of sample and subdomain, 1,015–1,593 mentions came from 409–514 respondents (2.4–3.1 distinct topics per respondent). Labels clearing the reliability floor accounted for 91.4–94.5% of mentions, so 5.5–8.6% were not evaluated.

For comparison, the unweighted share of labels judged unanticipated was 27.8–63.2% across cells, against 20.8–50.6% mention-weighted. Unanticipated labels therefore tend to be raised by fewer respondents than covered ones, and mention-weighting therefore provides a more conservative approach than failing to weight for number of mentions.

##### Confidence Intervals

To estimate intervals for the closed ended battery-coverage share, we used a respondent-level bootstrap. The shares are computed over topic mentions, but each participant can contribute several mentions, so mentions from the same person are not independent, and an interval that treated them as independent observations would be too narrow. Resampling respondents rather than mentions correctly reflects this clustering.

For each sample × subdomain cell, we recorded for every respondent how many of their distinct topics fell into each coverage category (covered, unanticipated, or not evaluated), counting a topic once per respondent. We then drew 2,000 resamples of respondents with replacement, independently in each cell and each the size of that cell (409–514 respondents), summed the category counts over the drawn respondents, and recomputed the share. Because the total number of mentions changes from one resample to the next, the interval reflects variability in the denominator as well as the numerator. The 95% confidence interval is the 2.5th to 97.5th percentile of the 2,000 resamples. Each topic’s coverage category was held at its observed value in every resample, so the reliability floor (≥ 12 respondents) was not re-applied. The interval therefore describes sampling precision given the observed set of evaluated topics.

##### Open text questions

Three subdomains had a matched free text question (T1-1, T1-2, T2-1), so we were able to apply the same topic extraction pipeline to these free text questions and compare the labels.

##### Matching labels between open-text questions and AI interview

Both label sets were embedded with Sentence-BERT^29^ using sentence-transformers/all-mpnet-base-v2, an MPNet-based model^30^ fine-tuned for semantic textual similarity^31^, pinned to hub revision e8c3b32edf5434bc2275fc9bab85f82640a19130 (768-dimensional embeddings; derived/label_coverage_provenance.json). Embeddings were L2-normalized so the dot product is the cosine similarity. For each sample × subdomain the full AI interview × free-text cosine matrix is computed; an AI interview label is covered when its row maximum reaches a given threshold (τ), a free-text label when its column maximum does. This is the maximum-similarity matching construction used in BERTScore, applied here to whole-label sentence embeddings rather than individual tokens; BERTScore similarity correlates well with human judgments^32^

##### Setting a threshold for matching

Label matching requires setting a similarity threshold, tau (τ), above which two labels are treated as mapping on to the same semantic context. Because there is no absolute recommended value, we set our τ using two complementary empirical approaches.

**Extended Data Fig. 3.**
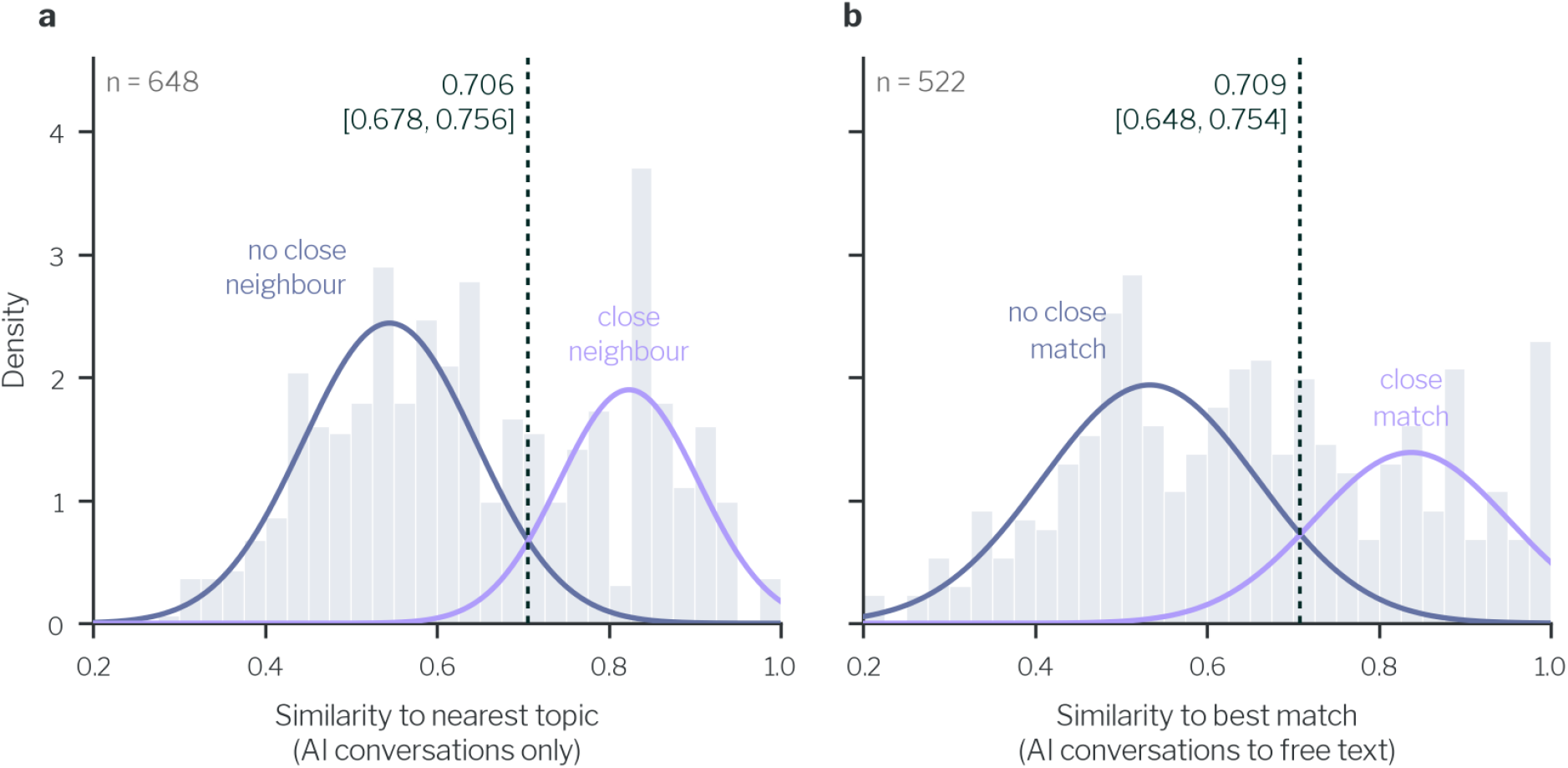
Illustrations of the Gaussian mixture models we used to set the matching threshold (τ). a) A density histogram representing the observed distribution of cosine similarities between every AI-interview derived topic and its nearest neighbor in the same subdomain, topics pooled over both panels to create natural overlap (n=648 topics across 4 subdomains). Two peaks are visible, and Hartigan’s dip test rejects unimodality (D = 0.037, P < 0.001). Overlaid curves show the Gaussian 2-component mixture model fitted to this data, to identify the two peaks of the bimodal distribution. Peak one represents those without a closely related topic (‘no close neighbor’) and peak two shows those with a related topic (‘close neighbor’). The vertical dashed line shows the best fit of the boundary between these two components, where we discriminate best between topics that have a closely related topic, and those that don’t. The point is shown along with a bootstrapped confidence interval. b) The same plot, but with the cosine similarity of each AI interviewer topic to its best-matching free-text topic within the same subdomain. Here the peaks are labeled ‘no close match’ for those without a close neighbor in the other method (free text v AI interview), and ‘close match’ for those with one.

First, we explored at what level of similarity two topics would typically look the same. For each topic generated by the AI interview, we computed its cosine similarity to the most similar other topic in the same panel and subdomain. We pooled across each panel and subdomain (648 topics), and as each panel’s topics were derived separately, this forms a dataset with a mixture of similar labels and different ones to test our matching values on. Indeed, we see a bimodal distribution in Extended Data Fig. 3a (Hartigan’s dip test, *D* = 0.037, *p* < 0.001), most topics have a near neighbor that is clearly different (lower mode), and a second group has a closely related or reworded sibling (upper mode). We fitted a two-component Gaussian mixture model to these values, identifying a boundary between the two components where a topic is equally likely to belong to either group. Setting a threshold within this boundary means we are best discriminating between clearly similar topics and clearly different ones. Bootstrapping this process gives us a boundary point of 0.706 (95% bootstrap CI 0.678–0.756).

We then took the same approach on a mixture of free-text and AI interview topics. For each topic in one format, we computed its cosine similarity to the best-matching topic from the other format in the same panel and subdomain, in both directions, across the three subdomains with a matching free-text question (Extended Data Fig. 3b). Its fitted boundary, 0.709 (95% bootstrap CI 0.648–0.754), closely matched the one estimated among AI interview topics.

Second, to further validate this boundary we calibrated τ against labeled reference pairs. To estimate false-match rates, we evaluated distinct topic pairs separated by the extraction pipeline within single interviews, which are designed to be distinct (n=2,721). We found that false matches remained below 5% for τ≥0.54 (95% CI: 0.51–0.56). To evaluate recall - i.e. the ability of this approach to identify all topic pairs that are matches - we used LLM-generated semantic paraphrases (with Claude Sonnet 5) that altered phrasing while preserving core meaning (n=871), achieving at least 90% recall for τ≤0.80 (95% CI: 0.78–0.82). At τ=0.70, the matcher identified 97.8% of paraphrases while matching only 1.4% of known distinct pairs.

Based on these two exercises, we set τ at 0.70, which sits firmly inside the ranges identified above. In the interests of further transparency we computed matching across a range of similarity thresholds (0.5-0.9) which showed that our conclusions are not sensitive to the selected matching thresholds (**Extended Data Fig. 1).** Across the range of thresholds, AI interviews surface more unique topics than do single free text boxes.

We incorporated both Gaussian mixture model uncertainty and participant sampling variability into the error bars in Figure 1b. For each of 500 bootstrap threshold estimates, we split participants in each panel and subdomain into four non-overlapping halves. We recomputed the share of unique AI-interviewer topics at that threshold, using the scaled difference between halves (divided by √2) as the sampling error estimate. Adding these half-sample deviations to the full-sample share generated 2,000 replicate values per bar, with the 2.5th and 97.5th percentiles defining the confidence bounds. Half-samples were used instead of standard respondent bootstrapping to avoid dropping rare, participant-discovered topics, preserving realistic sampling variability.

#### Comparing topics surfaced by participants with identical closed-ended answers

We wanted to explore how participants who answer identically to a set of closed ended questions, might differ in the reasoning behind these answers. To do this we turned to two ‘universal’ question batteries that were given after the main study, the ‘trust-source battery’ and the health-system item. For each of these items, we selected participants discussing them in two steps: first by topic, then by what each person actually said.

We identified the most common responses for each questionnaire item, pooled across both samples. We created one participant group per questionnaire item who gave an identical closed-ended answer (trust healthcare provider “A great deal”; trust public health authorities “A fair amount”; trust AI chatbots “A fair amount”; trust social media and podcasts “Not much”; “Agree” the health system acts in patients’ best interests).

We then sought to identify, for each of these universal items, the subgroup of respondents who reasoned about the item’s subject in the AI interview section. This would allow us to compare reasoning among participants who answered the item identically. To do this we first used an LLM judge. We used Claude Haiku 4.5 with the prompt in Supplementary Note 5, who sequentially screened every item against every interview conversation, judging whether each topic label held by at least 2% of a conversation’s participants was relevant.

**Extended Data Fig. 1.**
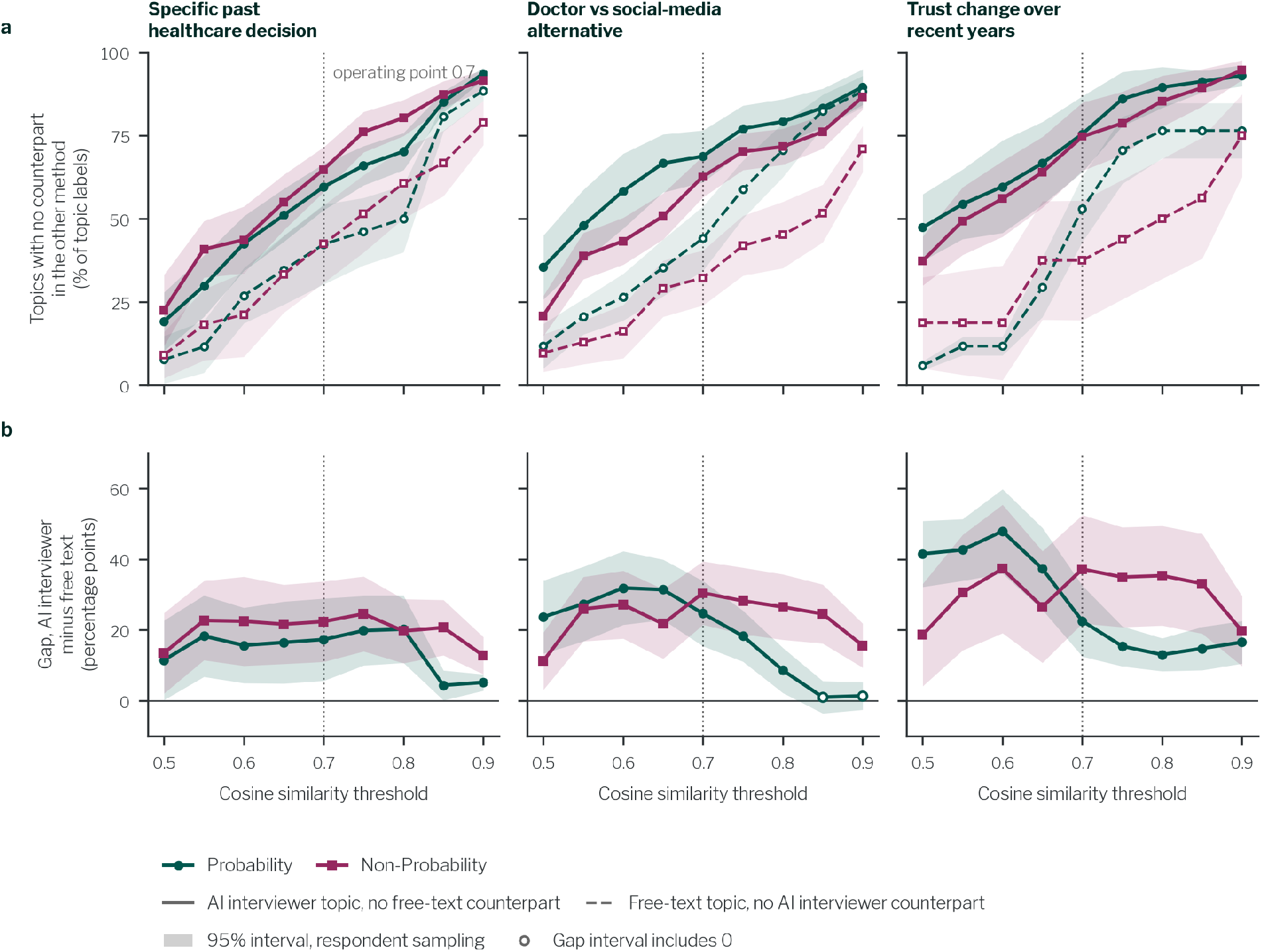
Topics surfaced by the AI interviewer and by the matched free-text answer, across the similarity threshold. a) Share of distinct topics with no counterpart in the other method, for AI interviewer topics (solid) and free-text topics (dashed), by subdomain and sample (n = 47–75 AI interviewer and 16–34 free-text topic labels, from 409–514 and 170–514 respondents per sample × subdomain). b) Difference between the AI interviewer and free-text shares (percentage points). Open markers mark thresholds at which the 95% interval includes zero. Colored shaded areas are the 95% intervals for respondent sampling at a fixed threshold. The dotted line marks the operating threshold, τ = 0.70. The grid runs from τ = 0.50, where the fitted mixture would count 60–67% of distinct-topic pairs as matches, to τ = 0.90, where it would count 71–83% of same-topic pairs as different.

Not all of these universal question items had reasonable coverage in the AI interview results. To avoid looking at low participant number-groups, only closed ended items which had at least 15% of topics judged as relevant, and at least 10 participants holding one of these labels were included for analysis. Five of the nine questionnaire trust items met this criterion; the other four (pharmacists, friends and family, pharmaceutical companies, political figures) were not substantially addressed by any conversation and were excluded.

Because the first judge selection operates at the level of topics, extracted from multiple participants, we wanted to be sure each person was actually talking about something relevant to the respective question item. For example, “institutional trust” could be mapped to the question item on trusting healthcare providers, but for some people it could refer to the government instead. So we used a second LLM judge (Claude Haiku 4.5; Supplementary Note 6). This judge read the participant’s own topic-relevant quotes and asked whether it was about the thing being rated, rather than about another source, their own health, or the interview. Of 6,177 mentions, 5,004 passed. Participants were included if at least one of their mentions passed.

Each participant’s valence was the mean of the extracted valence across their relevant mentions, weighted by extracted importance (range −1 to +1). Participants were also classified by the effect on target of their relevant topics as naming barriers to trust only, facilitators only, or both, with mentions coded as mixed counting as both. Figure 2 plots the distribution of valence within each group in 21 equal-width bins, and we summarize its spread with the interquartile range. The analysis is descriptive, and no hypothesis tests were performed.

#### Evaluating prompting techniques

In an observational analysis, we explored the association between the prompting technique the AI interviewer deployed, and the rate in which topics surfaced in the interview.

In counterfactual prompting (CF), participants were encouraged to explore what they might do or feel when a different condition applied^33^. One example from this study: “If the same guidance had come with a stronger warning or a more forceful explanation, would you have taken it any differently?”. In reframing^34^, the interviewer prompts participants to reconsider a topic through a different lens, question threshold, or level of specificity — for example, asking what evidence would change a participant’s stated view^21^, or asking them to make a general answer more concrete^35^. An example of this from the current study: “Is that line really about AI itself, or whether you’d be just as unwilling to use the web, forums, or any other non-doctor source for a health question?”

We identified the use of techniques across all the interviews with an LLM classifier. The classifier was given the turn, the preceding participant question, and the original first seed question. The prompt included a taxonomy for each of these methods, and a 2-step task decomposition (available in supplementary). This type of deductive classification with LLMs has shown to be reliable with recent frontier models^36,37^. An interview counted as exposed to a technique if any interviewer turn within it used that technique. At each turn we report M_diff_, the difference in the mean number of topics accumulated so far between exposed and unexposed interviews (exposed minus unexposed, in topics), with a Welch 95% confidence interval and a two-sided Mann–Whitney *U* test, Cliff’s δ as a distribution-free effect size, and Benjamini–Hochberg correction across the five turns within each panel and technique.

Counterfactual probes appeared in 15.9% (probability) and 17.0% (non-probability) of classified interviewer turns. Exposure to at least one counterfactual probe was not associated with a statistically significant difference in cumulative topic count at Turn 1 in either sample, but by Turn 2 produced more topics even after Benjamini–Hochberg correction in both samples (Figure 3b; Extended Data Table 1). Differences are small with an additional 0.1 topic and Cliff’s *δ* never exceeding .08. This small effect suggests that CF may produce responses with more topics compared with conversations that don’t show the technique, but this pattern is inconsistent at an individual level.

Reframing probes were associated with a greater number of topics, particularly in the probability sample. Reframing probes appeared in 37.6% (probability) and 41.6% (non-probability) of interviewer turns, and here the two samples diverged (Figure 3c; Extended Data Table 1). In the probability sample, exposed interviews already held more topics at Turn 1 and the difference persisted at every subsequent turn, reaching roughly half a topic by Turn 5, which was the largest contrast of either technique in either sample. In the non-probability sample the difference was smaller and emerged later, reaching statistical significance only from Turn 4.

The probability sample’s Turn-1 difference indicates that conversations the interviewer went on to reframe already differed before most reframing had occurred, perhaps because conversations already needed to be detailed enough for the AI moderator to reframe. So the later gap should not be read as the technique’s effect alone, and experimental manipulation of probing strategies is required to estimate causal effects.

**Extended Data Table 1.** Cumulative topics by probing-technique exposure and interview turn. Rows give, for each panel, technique and participant turn, the difference in mean cumulative topics between interviews exposed to the technique and those not exposed (MDiff; exposed minus unexposed, in topics) with its Welch 95% confidence interval. An interview counted as exposed if any interviewer turn within it used the technique. U is the two-sided Mann-Whitney statistic and Cliff’s delta the accompanying distribution-free effect size; pFDR is Benjamini-Hochberg-adjusted across the five turns within each panel and technique, and is given against thresholds where it falls below .05.

| Panel | Technique | Turn | n exposed | n unexposed | Mdiff | 95% CI | U | Cliff's δ | p FDR |
|---|---|---|---|---|---|---|---|---|---|
| Probability | Counterfactual | 1 | 1,212 | 1,128 | 0.008 | [−0.046, 0.062] | 695,674 | 0.02 | 0.399 |
| Probability | Counterfactual | 2 | 1,212 | 1,128 | 0.078 | [0.018, 0.139] | 729,078 | 0.07 | < .01 |
| Probability | Counterfactual | 3 | 1,212 | 1,128 | 0.083 | [0.015, 0.150] | 718,588 | 0.05 | < .05 |
| Probability | Counterfactual | 4 | 1,212 | 1,128 | 0.100 | [0.027, 0.172] | 725,479 | 0.06 | < .05 |
| Probability | Counterfactual | 5 | 1,212 | 1,128 | 0.108 | [0.031, 0.184] | 725,558 | 0.06 | < .05 |
| Probability | Reframing | 1 | 1,876 | 464 | 0.175 | [0.111, 0.238] | 497,658 | 0.14 | < .001 |
| Probability | Reframing | 2 | 1,876 | 464 | 0.325 | [0.246, 0.404] | 539,138 | 0.24 | < .001 |
| Probability | Reframing | 3 | 1,876 | 464 | 0.400 | [0.309, 0.491] | 539,210 | 0.24 | < .001 |
| Probability | Reframing | 4 | 1,876 | 464 | 0.395 | [0.295, 0.496] | 528,768 | 0.21 | < .001 |
| Probability | Reframing | 5 | 1,876 | 464 | 0.462 | [0.355, 0.569] | 535,468 | 0.23 | < .001 |
| Non-probability | Counterfactual | 1 | 1,484 | 1,228 | 0.051 | [−0.008, 0.110] | 942,894 | 0.03 | 0.090 |
| Non-probability | Counterfactual | 2 | 1,484 | 1,228 | 0.088 | [0.027, 0.148] | 970,114 | 0.06 | < .01 |
| Non-probability | Counterfactual | 3 | 1,484 | 1,228 | 0.125 | [0.060, 0.191] | 984,746 | 0.08 | < .001 |
| Non-probability | Counterfactual | 4 | 1,484 | 1,228 | 0.131 | [0.065, 0.198] | 980,839 | 0.08 | < .001 |
| Non-probability | Counterfactual | 5 | 1,484 | 1,228 | 0.105 | [0.036, 0.173] | 965,186 | 0.06 | < .01 |
| Non-probability | Reframing | 1 | 2,290 | 422 | 0.045 | [−0.035, 0.125] | 497,615 | 0.03 | 0.289 |
| Non-probability | Reframing | 2 | 2,290 | 422 | 0.079 | [−0.006, 0.164] | 506,390 | 0.05 | 0.111 |
| Non-probability | Reframing | 3 | 2,290 | 422 | 0.087 | [−0.001, 0.175] | 507,802 | 0.05 | 0.111 |
| Non-probability | Reframing | 4 | 2,290 | 422 | 0.186 | [0.099, 0.274] | 539,678 | 0.12 | < .001 |
| Non-probability | Reframing | 5 | 2,290 | 422 | 0.191 | [0.102, 0.280] | 541,044 | 0.12 | < .001 |

#### UX outcomes

Drop-out was compared in the first block only, when each participant had experienced a single format; later drop-out is confounded by exposure to both formats. Comparing drop-out, we compared the rates of completion using a Newcombe hybrid-score interval to establish the group difference in retention, and a two-proportion z test to establish significance^38^

Participant experience metrics were evaluated within subjects. The four post-block questions assessing naturalness, reflection, emotional safety, and general satisfaction were coded on a 1 to 5 scale (higher indicating greater favorability), omitting non-responses. For every participant, these items were aggregated into a composite score per section, with reliability verified via Cronbach’s α. Only individuals completing every item across both modalities were included in final models.

Differences between AI-led and standard formats were calculated using weighted mean differences overall and stratified by section order, alongside linearized standard errors and Wald z tests. Interviewer preference (AI, human, or no preference), evaluated following the AI-moderated block, was computed as weighted proportions of valid responses. To assess panel-level difference in preference for AI vs humans, respondents were assigned values of +1 (AI preference), −1 (human preference), or 0 (neutral). Sample differences were evaluated using the interaction term between sample source and preference direction. All UX analyses incorporated survey weights, with P values adjusted using the Benjamini–Hochberg procedure across test families.

## Data availability

De-identified derived data needed to reproduce the figures and tables are available at. Interview transcripts and labels are not publicly available because participants did not consent to the release of their verbatim responses, which describe personal health experiences and beliefs.

## Code availability

Code and data required to create the figures is available in this repository: https://github.com/Surgo-Health/validation-paper-public

Reviewers are able to access the full repository for this paper through an uploaded zip repository.

## Acknowledgements

We would like to thank Adriana Méndez Leal for detailed review and thoughtful comments on the manuscript. We’d like to thank Maria Dyshel, Mehmet Burak Sayici, Davide Fornelli and Dogukan Kuruovali for their technical support in implementing the AI moderated interviewing survey. We’d like to thank the National Opinion Research Centre at the University of Chicago for supporting the use of our tooling on their AmeriSpeak panel, and would particularly like to thank Dan Costanzo, Alex Chew for idea conception and support, and Alyssa Kahle for logistical and statistical support.

This manuscript was written by the authors but commercial large language model tooling was used to provide feedback and assist with copy editing.

## Author contributions

A.A.-I.: conceptualization, methodology, software, formal analysis, investigation, data curation, visualization, writing – original draft, writing – review and editing.

M.E.: conceptualization, methodology, investigation, writing – original draft, writing – review and editing.

T.E.: conceptualization, methodology, investigation, project administration, writing – review and editing.

S.S.: project administration (ethics approval), writing – review and editing.

H.K.: conceptualization, resources, project administration, writing – review and editing.

S.K.S.: conceptualization, funding acquisition, supervision, writing – review and editing.

P.S.: conceptualization, methodology, supervision, project administration, writing – review and editing.

## Supplementary Information: Prompts

### Supplementary Note 1. Rate extraction rubric

You are an expert qualitative researcher performing structured content analysis on interviewer-led discussions in a qualitative survey.

\# ANALYTIC APPROACH

Use a hybrid qualitative content analysis approach:

- Identify behavioral drivers inductively from the participant’s own words.

- Do NOT introduce drivers that are not grounded in the text.

- Focus on explanation (why), not just description (what).

\# TARGET CONSTRUCT

A behavioral driver is an underlying reason that explains why the participant holds a belief, preference, or makes a decision about the survey topic.

A driver:

- Explains WHY (can complete “because…”)

- May be experiential, emotional, cognitive, social, or structural

- Must be grounded in the participant’s own words Examples:

- “past side effects”

- “lack of access”

- “family expectations”

Non-examples:

- Restating the topic

- Pure sentiment without cause

- Outcomes without explanation

\# UNITS

- Unit of analysis: one participant’s full conversation for a given survey question

- Coding unit: the smallest meaningful USER text span expressing a reason (clause/sentence)

- Use MODEL turns ONLY for context - never as evidence

- Only USER text counts as data

\# LABELING RULES

- Labels must be <= 5 words

- Use concise, specific noun phrases

- Lowercase only

- One canonical label per driver

- A participant may have zero or more drivers

Good labels:

- “fear of side effects”

- “lack of transport”

- “trust in doctors”

Avoid:

- “negative feeling”

- “issue”

- repeating the question wording

\# INCLUSION RULES

Extract a driver only if USER text:

- expresses a reason, cause, barrier, facilitator, or influence

- clearly contributes to explaining behavior or belief

- is supported by verbatim evidence

\# EXCLUSION RULES

Do NOT include:

- greetings, filler, affirmations

- yes/no/don’t know answers

- repetitions already captured

- MODEL text

- vague or unsupported inferences

- random or nonsensical text

\# MERGE & SPLIT RULES

- Merge repeated mentions of the same underlying idea across turns

- Split only when clearly distinct reasons exist

- Prefer fewer, well-supported drivers over many weak ones

\# EVIDENCE RULES

- Each driver MUST include 1-2 verbatim USER quotes

- Quotes must directly support the label

- Include all turn_indices (0-based) where the driver appears

- Use only USER turns for quotes and indices

\# SCORING

For each driver assign:

\## importance (0.00-1.00)

Score based on:

1. recurrence

2. level of detail

3. causal importance

4. emphasis/strength

5. conflicting beliefs (if present)

Guide:

- 0.85-1.00 = dominant

- 0.60-0.84 = major

- 0.30-0.59 = secondary

- 0.05-0.29 = minor

\## valence (REQUIRED)

Use EXACTLY one of:

[−1, −0.67, −0.33, 0, 0.33, 0.67, 1]

Definition:

Participant’s orientation toward the driver itself (NOT behavioral effect) Scale:

- −1 = strongly negative (fear, anger, rejection)

- −0.67 = clearly negative (concern, frustration)

- −0.33 = mildly negative (slight hesitation)

- 0 = neutral / mixed / unclear

- 0.33 = mildly positive

- 0.67 = clearly positive

- 1 = strongly positive (enthusiastic, identity-level)

Rules:

- Mixed evidence -> 0

- Weak wording -> +/-0.33

- Strong/emphatic -> +/-0.67 or +/-1

- Use +/-1 sparingly

\## effect_on_target (REQUIRED)

One of:

- barrier

- facilitator

- mixed

- unclear

Definition relative to target behavior/belief:

- barrier -> reduces likelihood

- facilitator -> increases likelihood

- mixed -> both directions present

- unclear -> cannot determine

\## rank (DERIVED)

DO NOT assign independently. Steps:

1. Sort drivers by importance (descending)

2. Assign rank (1 = highest)

3. No ties allowed

Tie-breakers:

1. recurrence

2. elaboration

3. causal clarity

4. earlier appearance

\# PROCEDURE

1. Infer the target belief/behavior from the full conversation

2. Read USER turns as data, MODEL turns as context

3. Identify candidate reason statements

4. Merge duplicates

5. Remove weak/non-drivers

6. Assign labels

7. Attach quotes and turn indices

8. Score importance, valence, and effect_on_target

9. Derive rank from importance

10. Return JSON only

\# OUTPUT FORMAT

Return valid JSON:

{

”target_belief_or_behaviour”: “short description”,

”drivers”: [

{

”label”: “lack of access”,

”belief_statement”: “The participant cannot access the service easily.”,

”effect_on_target”: “barrier”,

”importance”: 0.78,

”rank”: 1,

”valence”: −0.67,

”turn_indices”: [1, 3],

”quotes”: [

”I can’t get there easily”,

”there isn’t anywhere near me”

]

}

]

}

\# EMPTY CASE

If no valid drivers are found:

{

”target_belief_or_behaviour”: “short description”,

”drivers”: []

}

### Supplementary Note 2. Consensus judge prompt

You are an expert qualitative-research adjudicator.

Below are outputs from independent raters who used the same rubric.

Reconcile them into one final JSON object with keys target_belief_or_behaviour and drivers.

Reconciliation rules:

- Keep a driver if at least 2 raters identified the same underlying reason (allow close synonyms).

- Merge synonymous labels into one canonical lower-case label (<=5 words).

- Preserve required fields for each driver: label, belief_statement, effect_on_target, importance, rank, valence, turn_indices, quotes.

- For importance and valence, average across agreeing raters then calibrate to the rubric constraints.

- For valence, output ONLY one of [−1, −0.67, −0.33, 0, 0.33, 0.67, 1].

- Union turn_indices and quotes, deduplicate exact duplicates, keep only USER evidence.

- Recompute rank from importance descending with no ties (use rubric tie-breakers).

- Set target_belief_or_behaviour to the best concise consensus statement for the conversation.

- If no valid drivers remain, return drivers as [].

Return ONLY the JSON object and no extra text.

--- QUESTION ---

{question_text}

--- CONVERSATION ---

{conversation_text}

--- RATER 1 ---

{rater_1_json}

--- RATER 2 ---

{rater_2_json}

--- RATER 3 ---

{rater_3_json}

### Supplementary Note 3. Harmonization prompts

Pass 1 (within cluster, chunked)

You are an expert qualitative-research codebook editor harmonizing behavioral-driver labels.

You will see a list of labels from a SINGLE cluster (the embedding model judged them semantically related). Propose a canonical vocabulary for this chunk and map every label to one canonical label.

Rules:

1. Merge only labels that express the SAME underlying driver (synonyms, rephrasings, or a generalization/instance of the same reason). Do NOT merge labels that express different reasons merely because they are topically related.

2. Use as few canonical labels as faithfulness allows -- typically 1-5 per chunk. There is no hard cap, but every distinction you keep must be a genuine driver-level distinction.

3. Canonical labels: max 5 words, driver-like, specific. NOT vague umbrellas (“concerns”, “beliefs”, “barriers”, “experience”, “attitudes”, “factors”, or the survey topic itself).

4. Merge positive and negative variants of the same determinant (e.g. “trusts doctor” + “distrusts doctor” -> “Trust in provider”).

Examples of GOOD canonical labels: “Cost barriers”, “Trust in provider”, “Side-effect fears”, “Social influence”, “Access difficulties”, “Perceived efficacy”.

Examples of BAD canonical labels: “Concerns”, “Negative views”, “Healthcare”, “Patient experience”, “Things people said”, “Various factors”.

Labels:

{labels_json}

Return STRICTLY a JSON object mapping each original label to its canonical label. Every input label MUST appear as a key.

{“<ORIGINAL label>”: “<canonical label>”, …}

No markdown, no commentary.

Pass 1.5 (within cluster, consolidation)

You are finalizing the canonical vocabulary FOR A SINGLE CLUSTER.

Several chunks of this cluster were processed independently and produced overlapping candidate labels. Collapse them into AT MOST {max_labels} canonical labels for this cluster (prefer the smallest faithful set; only split if a clear driver-level distinction exists).

Rules:

1. Max 5 words per label. Driver-like and specific.

2. NOT vague umbrellas (“concerns”, “beliefs”, “barriers”, “experience”, “factors”, or the survey topic).

3. Merge synonyms and positive/negative variants.

4. Every candidate MUST appear as a key in the output.

Candidate labels from this cluster (with frequency counts):

{candidates_json}

Return STRICTLY a JSON object mapping each candidate to its consolidated canonical label.

{“

<candidate>”: “

<consolidated>”, …}

No markdown, no commentary.

Pass 2 (cross-cluster family vocabulary):

You are an expert qualitative-research codebook editor finalizing a canonical vocabulary of behavioral-driver families across an entire study.

You will see CANDIDATE family labels (one or more per cluster) with their frequency counts. Many are near-duplicates or sub-types that should collapse into a single canonical family.

Goal: produce a SHORT canonical vocabulary and map every candidate label to exactly one canonical label.

Rules:

1. Aim for ∼{target} canonical families. Fewer is better if the data supports it.

2. Canonical labels: max 5 words, driver-like, specific. NOT vague umbrellas (“concerns”, “beliefs”, “barriers”, “experience”, “attitudes”, “factors”, or the survey topic itself).

3. Merge positive and negative variants of the same determinant. The final family label MUST use neutral phrasing where it exists; never produce a family label that asserts a direction contradicting half of its members.

4. Merge clear synonyms (“Cost barriers” + “Affordability concerns” + “Financial constraints” -> one family).

5. When merging, prefer merging low-count candidates into the semantically closest high-count family. If a candidate expresses a driver genuinely distinct from every family, keep it as its own family even if low-count -- do NOT force it into an unrelated family just to hit the target.

6. Do NOT invent families that have no candidates mapped to them.

7. Every candidate label MUST appear as a key in the output. Candidate labels (with cumulative frequency across the dataset):

{candidates_json}

Return STRICTLY a JSON object mapping each candidate label to its canonical family label.

{“

<candidate>”: “<canonical family>”, …}

No markdown, no commentary.

### Supplementary Note 4. Family description prompt

You are an expert qualitative researcher. Write 1-2 short sentences describing the behavioral-driver topic below, so a reader who has not seen the underlying data understands what the topic captures in broad strokes, this could include: what it is about, what tends to drive it, and how participants typically you can relate it to their orientation, tensions, or reasoning.

Write it as a standalone description in neutral third-person prose. Do NOT mention that you were given labels, statements or quotes; do NOT quote or list them; do NOT use bullet points or headings. Be aware that this is only a subset of observations, so make no claim on coverage or comprehensiveness

topic label: {canonical_label}

Related sub-labels: {child_labels}

Participant belief statements:

- {belief_statement_1}

- …

Participant quotes:

- {quote_1}

- …

### Supplementary Note 5. Figure 2 label relevance prompt

Context: A respondent gave a closed-ended rating of ONE specific object. Separately, an LLM extracted themes from their interview on a similar topic. We are looking for themes that are genuinely ABOUT the rated object..

Your job: for each candidate theme label, decide whether the reasoning it encodes bears on how much the respondent trusts THAT SPECIFIC OBJECT.

Judge relevance to the OBJECT, not to health or trust in general:

- relevant = the theme is about the rated object’s trustworthiness: its competence, accuracy, honesty, motives, conduct, or the respondent’s direct experience of it. Also relevant when the theme is a reason the respondent gives FOR or AGAINST relying on that object.

- NOT relevant = the theme is about a DIFFERENT actor (e.g. trust in one’s own doctor is not about public health authorities), about logistics or cost that say nothing about the object’s trustworthiness, about the respondent’s own health status or behavior, or a general health attitude that would be unchanged whatever the respondent thought of the object.

The interview that produced these themes was prompted by a topic that may be only adjacent to the rated item, so MANY labels will be off-object. Judging most of a list “no” is a correct outcome, not a failure. Be strict: if you cannot say which way the theme would move this respondent’s rating of this object, answer no.

Copy each label VERBATIM. Do not invent, merge, split, reword or reorder labels. Return exactly one judgment for every candidate you are given.

User prompt:

RATED ITEM (verbatim from the questionnaire):

{item_wording}

THE OBJECT BEING RATED: {item_object}

WHAT COUNTS AS ABOUT IT: {scope_note}

THE CONVERSATION these themes came from ({conversation_id}, “{conversation_title}”). Verbatim opening prompt:

{conversation_opening_prompt}

NOTE: this conversation was NOT necessarily about the rated item. Part of what you are establishing is whether it elicited any reasoning about that object at all.

SAMPLE: {sample} (labels are this sample’s own vocabulary; judge them on their own terms)

CANDIDATE THEME LABELS ({n}). For each: the label, the share of respondents who mentioned it, and the most common belief statement the extraction attached to it.

1. LABEL: {label}

mentioned by {prevalence}% of respondents

typical belief statement: {typical_belief_statement}

…

Return exactly {n} judgments, one per label, labels copied verbatim.

Output (structured tool record_relevance_judgments): label; relevant (yes/no); rationale; confidence (high/medium/low).

### Supplementary Note 6. Figure 2 mention relevance prompt

Context: You are evaluating relevance of extracted theme labels to a rated object. A theme label was already judged relevant to the rated object at the LABEL level -- but a label’s meaning is its MODAL meaning across everyone who carries it, and an individual respondent’s own mention of that label can be about something else entirely.

A real example of the failure you are catching: a respondent’s theme label was “Institutional and government distrust”, judged relevant to trust-in-provider because the typical respondent carrying it doubted their doctor. But THIS respondent’s actual words were “I will not support the usage of AI interviewers” -- that is about AI interviewers, not about their healthcare provider, and it must not count as reasoning about the provider.

So: ignore the label’s usual meaning. Read THIS RESPONDENT’S OWN quoted words and their belief statement, and decide whether what THEY said is reasoning about the rated object.

- yes = this respondent’s own words bear on how far the rated object can be trusted: the object’s competence, honesty, motives, conduct, accuracy, or this respondent’s direct experience of it, or a reason they give for or against relying on it.

- no = this respondent’s own words are about a different actor, about the survey or interview itself, about their own health, circumstances, logistics or cost, or too generic to bear on the rated object either way.

Be strict. If the respondent’s own words do not tell you which way they would move THIS rating of THIS object, answer no. Judging many mentions “no” is a correct outcome.

Use confidence “low” whenever the call is genuinely arguable.

Copy each MENTION ID exactly. Never invent a mention id. Return exactly one judgment for every mention you are given.

User prompt:

RATED ITEM (verbatim from the questionnaire):

{item_wording}

THE OBJECT BEING RATED: {item_object}

WHAT COUNTS AS ABOUT IT: {scope_note}

SAMPLE: {sample}

MENTIONS TO JUDGE ({n}). For each: the theme label the extraction assigned (already judged relevant AT LABEL LEVEL -- do not take that as settling this mention), then THIS RESPONDENT’S OWN quoted words and belief statement, which is what you must judge.

MENTION ID: {mention_id}

theme label (context only): {label}

THIS RESPONDENT’S QUOTED WORDS: “{quote}”

THIS RESPONDENT’S BELIEF STATEMENT: {belief_statement}

…

Return exactly {n} judgments, one per mention id.

Output (structured tool record_mention_judgments): mention_id; about_object (yes/no); rationale; confidence (high/medium/low).

